# Health and Economic Benefits of Air Quality Improvements in France through Net-Zero Transition Scenarios by 2050

**DOI:** 10.64898/2026.05.27.26354123

**Authors:** Ayushi Sharma, Alicia Gressent, Elsa Real, Khanh Ninh Nguyen, Magali Corso, Mathilde Pascal, Sylvia Medina, Vérène Wagner, Rémy Slama, Augustin Colette, Kévin Jean

**Affiliations:** Biology department of the Ecole Normale Supérieure (IBENS), CNRS, INSERM, Paris, France; Paris Reasearch in Health, Environment, Climate (PARSEC), Ecole Normale Supérieure (ENS), INSERM, Paris, France; Institut National de l’Environnement Industriel et des RISques (INERIS), Parc Technologique ALATA, BP 2, 60550 Verneuil-en-Halatte, France; Santé publique France (SpF), 12, rue du Val d’Osne, 94415, Saint-Maurice Cedex, France

**Keywords:** Air pollution, Mortality, Climate mitigation, Net-Zero, Climate-Policy, Health Economic co-benefits

## Abstract

**Background:** Climate mitigation policies just with air quality improvements can deliver substantial co-benefits through reductions in greenhouse gas emissions and air pollutant concentrations. Mitigation strategies range from sufficiency to technology-driven transitions, yet linked co-benefits remain poorly understood. This study evaluates PM_2.5_ and NO_2_ exposure changes and associated co-benefits using France’s energy-transition scenarios.

**Methods:** Emission projections were incorporated to the CHIMERE chemistry-transport model to estimate PM_2.5_ and NO_2_ concentrations for 2030 and 2050. Health impacts were assessed using disease-specific cessation-lag assumptions relative to 2019, covering premature mortality, morbidity, DALYs, and economic benefits across nine outcomes (hypertension, lung cancer, ischaemic heart disease, stroke, COPD, type-2 diabetes, acute lower respiratory infections, and asthma in children and adults).

**Findings:** Population exposure in continental France is projected to decline by 23% and 45% for PM_2.5_ and NO_2_ by 2030, rising to 40% and 70% by 2050. Health gains are substantial and broadly consistent across all four scenarios, with modest differences between sufficiency-oriented and technology-driven pathways. Under delayed-impact assumptions, which accounts for the latency between exposure reduction and health response, avoided premature deaths reached 8,800-9,300 for PM_2.5_ and 11,000-12,800 for NO_2_ in 2030, rising to 21,300-22,100 and 24,500-26,200 by 2050. Avoided morbidity cases grew from 51,100–55,200 in 2030 to 84,000-87,800 by 2050, with total DALYs averted increasing from 278,000-310,000 to 427,000-450,000 over the same period. Economic benefits scaled accordingly, direct medical cost savings reached €1.0-1.1 billion/year by 2050, with intangible cost savings of €41-43 billion with PM_2.5_ and €36-39 billion with NO_2_ reductions.

**Conclusion:** Net-zero transition delivers substantial, progressive health and economic co-benefits that are robust across diverse policy pathways. Rather than sectoral composition, commitment to decarbonisation itself drives air quality improvements and population health gains. These findings support integrating co-benefits into climate policy frameworks to strengthen the evidence base for ambitious mitigation action.

**Highlights:**

- Net-zero policies can deliver large air-quality co-benefits; here, chemistry-transport modeling coupled with emission projections quantifies these benefits across four contrasting French decarbonisation pathways for 2030 and 2050.
- Health and economic co-benefits are large and robust across all four contrasting scenarios, with modest differences between sufficiency-oriented and technology-driven pathways.
- By 2050, PM_2.5_ and NO_2_ concentrations decline by 38-40% and 70-75%, averting 20,000-22,000 and 24,500-26,200 attributable premature deaths annually.
- Avoided morbidity reaches 84,000-87,800 cases/year and DALYs averted reach 427,000-450,000/year in 2050, growing substantially from 2030 estimates.
- Monetization of health impacts results in, direct medical savings of €1 billion/year in 2050 and intangible cost savings of €43 billion (PM_2.5_) and €38 billion (NO_2_) per year.

## 1 Introduction

Global commitment to net-zero (NZ) greenhouse gas (GHG) emissions, driven by the Paris Agreement’s goal to limit warming below 2*^◦^*C, has led to widespread national targets and strengthened climate policy.^1^ The IPCC estimates that GHG emissions must fall by about 43% by 2030 to meet temperature goals, requiring immediate, deep, and sustained action.^2^ Current Nationally Determined Contributions (NDCs) are insufficient for the 1.5*^◦^*C target, but the IPCC Sixth Assessment Report notes that decarbonisation also brings near-term health benefits, especially via improved air quality, as many GHG sources overlap with air pollutant sources.^2^

Air pollution remains a major public health issue in Europe despite reduced anthropogenic emissions. The EEA *Air Quality Status in Europe 2026* reports that although NO_2_ and PM_2.5_ levels far exceed WHO guidelines, 92.5% of monitoring stations surpass the PM_2.5_ annual guideline, and all countries exceed the NO_2_ WHO guideline value, exposing over 90% of Europeans to harmful PM_2.5_ and NO_2_ concentrations. Note that while the revised Ambient Air Quality Directive (AAQD 2024) tightens EU limit values, these remain above WHO guidelines and are not yet in force. In France, air pollution is a major public-health concern. Although the electricity system is largely decarbonised via nuclear power, sectors such as transport, industry, buildings, and agriculture still require rapid emissions cuts to meet the 2050 carbon-neutrality target. Between 2016 and 2019, Santé publique France attributed about 40,000 premature deaths annually to PM_2.5_ and 7,000 to NO_2_, with some overlap between the pollutants.^3^ In Paris, PM_2.5_ and NO_2_ fell by 32% from 2010 to 2024,^4^ yet further reductions are needed across multiple sectors: residential wood heating is the leading source of primary PM_2.5_, transport is a major NO_2_ source (31% of French GHG emissions),^5^ and industry contributes significantly to both pollutants.

The French Agency for Ecological Transition (ADEME) developed national scenarios aligned with international carbon neutrality targets for 2050.^6^ These scenarios offer potential co-benefits including improved air quality, health benefits, increased physical activity, and dietary gains.^7–10^ In addition to technological changes, behavioural measures such as increased walking, cycling, and public transport reduce emissions. The industrial sector is not explicitly included and falls outside this study’s scope. Meanwhile, the ADEME scenarios do not explicitly address the industrial sector, which therefore falls outside the scope of the present study.

Finally, a recent systematic review indicates that existing research on climate mitigation co-benefits predominantly focuses on PM_2.5_-attributable mortality, leaving alternative pollutants, morbidity, and diverse health-economic approaches under-evaluated.^8^ Furthermore, integrated health-economic assessments comparing prospective contrasting NZ scenarios are scarce. This evidence highlights a significant gap in the literature regarding prospective NZ transition trajectories, despite their known co-benefits. Particularly, in France, to date, no national-level studies have conducted fine-scale evaluations of prospective air quality improvements and their co-benefits. Addressing this evidence gap is essential for robust, policy-relevant prioritization. Therefore, this study quantifies mortality, morbidity and economic co-benefits associated with projected air-quality improvements across the multiple NZ scenarios for continental France.

## 2 Methodology

### 2.1 ADEME’s *Transition(s) 2050* Scenarios

The French Ecological Transition Agency (ADEME) proposed four heuristic *Transition(s) 2050* scenario to achieve carbon neutrality in continental France.^11^ These scenarios share the same macroeconomic, demographic, and climate assumptions (+2.1°C in 2100) but differ in societal and technological choices across energy, mobility, housing, and agri-food. All scenarios substantially reduce GHGs and key air pollutants, although the mix of levers differs. The emission changes described are taken directly from the ADEME scenario inputs and are taken into consideration in the present modelling work Table 1.

**Table 1:**
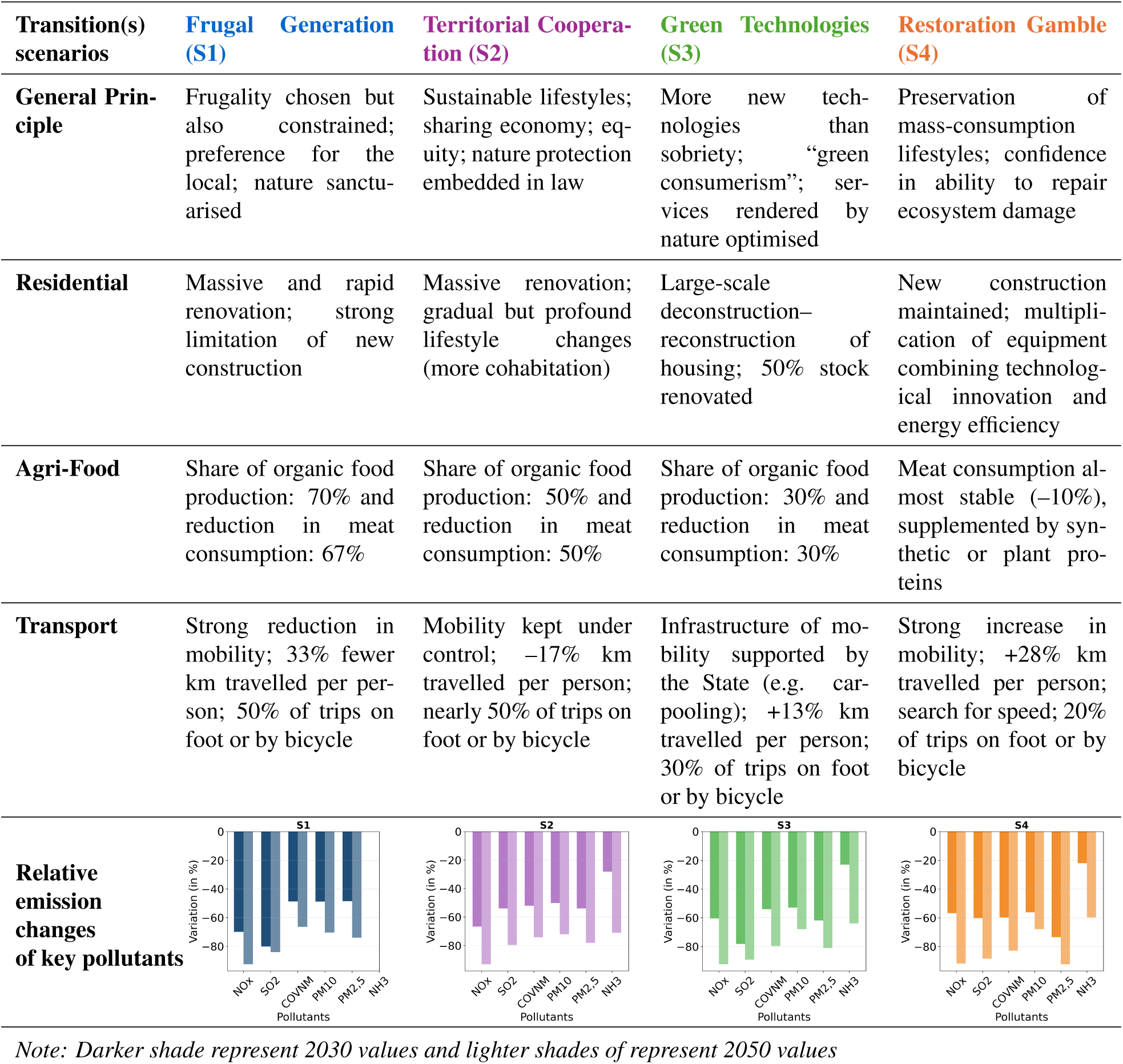
Overview of sectoral narrative of ADEME’s Transition(s) 2050 scenarios. ^11^

Sufficiency scenarios (S1, S2) achieve larger cuts in energy and transport pollutants (PM_2.5_, NO*_x_*, SO_2_, VOCs) via lower demand, building retrofits, and shifts to organic diets, which also reduce NH_3_ from agriculture. S1 represents the most ambitious sufficiency pathway, while S2 combines sufficiency with stronger regulatory standards. Technology-focused scenarios (S3, S4) prioritise efficiency and innovation, with higher travel demand and smaller dietary shifts, leading to smaller reductions in transport and agricultural pollutants.S3 assumes a more cooperative governance context, whereas S4 reflects a more fragmented, market-led transition. Yet, pollutant trends remain similar, indicating varied policy mixes can yield comparable air-quality gains. Detailed sectoral-specific assumptions are given in Annex-I.

### 2.2 Air Quality Modelling

We performed the Chemical Transport Modelling (CTM) with CHIMERE,^12^ a regional-scale air quality model that simulates the chemistry, transport, and deposition of atmospheric pollutants by integrating meteorological inputs, emission inventories, boundary conditions, and chemical mechanisms. The model captures how changes in precursor emissions affect pollutant concentrations through a process based on spatial grid discretization simulation. ADEME’s future 2030 and 2050 scenarios and the 2019 baseline were modelled at a resolution of about 3x3 *km*^2^ and subsequently downscaled and bias corrected on a basis of a reanalysis^13^ at a spatial resolution of 2 km × 2km and 1 km × 1 km, for PM_2.5_ and NO_2_, respectively. The 2019 baseline simulations used national emission inventories from CITEPA (OMINEA 2022), selected for consistency with the ADEME scenario methodology; the same methodological choices including exclusion of condensable emissions from domestic wood heating were applied across all runs.

ADEME’s future emissions contained gaps requiring filling prior to preprocessing, as complete data were needed for all relevant pollutants (PM_2.5_, PM_10_, NO*_x_*, SO*_x_*, NMVOCs, and NH_3_) and GNFR sub-sectors across all four scenarios. The projected inventories therefore combine ADEME emissions where available with CITEPA 2030 and 2050 estimates from the third French National Low-Carbon Strategy (SNBC3), the latter covering sectors such as industry for which ADEME provided no projections. Complete inventories were then preprocessed through spatialization (CAMS-REG),^14^ temporalization (CAMS-TEMPO),^15^ and speciation (emiSURF).^12^ Concentration changes were calculated relative to the 2019 baseline, whose modelled outputs were bias-corrected via a spatially explicit adjustment combining modelled concentrations with permanent monitoring network measurements (following French National Institute for Industrial Environment and Risks, INERIS, ambient air pollutant map methodology).^13^ Gridded concentrations were spatially matched to IRIS units using area-weighted averaging, then aggregated to commune level by population weighting to ensure exposure estimates reflect where people live (Annex-IV).

### 2.3 Health and Demographic Data

National all-cause mortality data and demographic projections for 2030 and 2050 were sourced from the French National Institute of Statistics and Economic Studies (INSEE), consistent with those used by ADEME for *Transition(s) 2050* scenarios. Prospective national population data were downscaled to IRIS level for age groups using proportional allocation, for each age group, the projected national change from the 2019 baseline to the target year (2030 or 2050) was applied to the corresponding IRIS-level baseline, preserving local demographic heterogeneity and national consistency. Mortality baselines thus vary across projection years according to anticipated demographic change. Methodological details and key assumptions are in Annex-II and III.

For morbidity endpoints, municipality-level average cause-specific incidence (2016–2019) was obtained from the French National Health Data System (SNDS). Disease selection was based on established causal links with air pollution exposure,^16^ covering nine diseases: (1) *cardiovascular:* stroke, acute myocardial infarction (AMI), hypertension; (2) *respiratory:* lung cancer, COPD, asthma (children and adults), acute lower respiratory infections excluding influenza (ALRI); and (3) *metabolic:* type-2 diabetes. Age-specific concentration–response functions (relative risks) for PM_2.5_ and NO_2_ were sourced from meta-analyses and recent report of WHO^16,17^ (Table 2). Unlike mortality, morbidity incidence rates were held constant across projection years due to the absence of comparable prospective estimates at the required spatial resolution.

**Table 2:** Input data used in the air-pollution health impact assessment.

| Health outcome<br>(ICD-10 codes) | Age range | Disability weight | Duration (years) | RR (95% CI)<br>per 10 $\mu\text{g}/\text{m}^3$<br>$\text{PM}_{2.5}$ | RR (95% CI)<br>per 10 $\mu\text{g}/\text{m}^3$<br>$\text{NO}_2$ | RR Reference |
| --- | --- | --- | --- | --- | --- | --- |
| All-cause mortality | 30+ | – | – | 1.10 (1.06–1.13),<br>(5–70 $\mu\text{g}/\text{m}^3$) | 1.05 (1.03–1.07),<br>(10–130 $\mu\text{g}/\text{m}^3$) | HRAPIE-II<sup>18</sup> |
| Hypertension<br>(I10–I11) | 18–99 | 0.08 | 15.6 | 1.17 (1.05–1.30) | – | Qin et al., 2017<sup>19</sup> |
| Lung cancer<br>(C34) | 35–99 | 0.13 | 2.12 | 1.16 (1.10–1.23) | – | Yu et al., 2021<sup>20</sup> |
| Asthma in children<br>(J45) | 0–18 | 0.04 | 16.7 | 1.34 (1.10–1.63) | 1.10 (1.05–1.18) | Khreis et al.,<br>2017<sup>21</sup> |
| Asthma in adults<br>(J45) | 18–99 | 0.04 | 16.7 | – | 1.10 (1.01–1.21) | HRAPIE-II<sup>18</sup> |
| COPD (J41–J44) | 40–99 | 0.03 | 14.4 | 1.18 (1.13–1.23) | – | Park et al.,<br>2021<sup>22</sup> |
| ALRI in children<br>(J12–J18, J20–<br>J22) | 0–12 | 0.07 | 0.02 | – | 1.09 (1.03–1.16) | HRAPIE-II<sup>18</sup> |
| Stroke (I60–I64) | 35–99 | 0.15 | 9.70 | 1.16 (1.12–1.20) | – | Yuan et al.,<br>2019<sup>23</sup> |
| AMI events<br>(I21–I22) | 30–99 | 0.03 | 6.90 | 1.13 (1.05–1.22) | – | Zhu et al., 2021<sup>24</sup> |
| Type 2 diabetes<br>(E11–E14) | 45–99 | 0.07 | 24.5 | 1.10 (1.03–1.18) | – | Yang et al.,<br>2020<sup>25</sup> |
| **Sensitivity analysis** |  |  |  |  |  |  |
| All-cause mortality | 30+ | – | – | 1.15 (1.05–1.25) | 1.02 (1.01–1.04) | $\text{PM}_{2.5}$: Pascal, 2016;<sup>26</sup><br>$\text{NO}_2$: COMEAP<br>2010<sup>17</sup> |

Note: ALRI: acute lower respiratory infections; AMI: acute myocardial infarction; COPD: chronic obstructive pulmonary disease

### 2.4 Health Impact Assessment (HIA)

Health impacts were assessed with age-stratified analysis for 2030 and 2050, linking projected changes in PM_2.5_ and NO_2_ to all-cause mortality and cause-specific morbidity. Annual concentration changes (Δ*C*) in each commune were computed relative to 2019 as population-weighted means and combined with concentration–response functions (Table 2) for each endpoint. Premature deaths averted due to exposure reductions were estimated by age group and commune as:

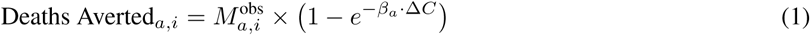

where 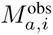 denotes the observed all-cause mortality in age group *a* and commune *i* during the reference year, obtained from vital statistics. Here, *β_a_* is the age- and endpoint-specific concentration–response coefficient, computed from the selected relative risk (RR) per 10 µg.m^-3^ increment. Relative risks (RR) were assumed log-linear. Age (*a*) and commune-specific (*i*) attributable fractions (AF) were also estimated, representing the proportion of mortality in the exposed population attributable to the pollutant. Under the log-linear assumption, the AF is computed as:

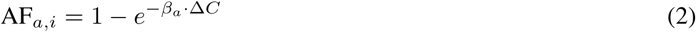

which is equivalent to the epidemiological definition AF*_a,i_* = (RR*_a,i_* − 1)*/*RR*_a,i_*. To contextualize the projected gains across the scenarios, a counterfactual reference threshold of 5µg.m^-3^ and 10µg.m^-3^ for PM_2.5_ and NO_2_, was applied on baseline (2019), based on WHO Air Quality Guidelines. Additionally, a more stringent theoretical minimum-risk counterfactual of 3µg.m^-3^ and 1µg.m^-3^ were also applied on 2019 reference concentrations, to illustrate the upper bound of potential co-benefits.

Years of Life Lost (YLL) averted were estimated by multiplying avoided deaths by remaining life expectancy at each age, then summed across ages and communes. Morbidity impacts covered nine incident diseases; for each age group and commune, avoided incident cases used the AF formula above, with mortality replaced by age-specific incidence rates.

Disability-adjusted life years (DALYs) were calculated by adding YLL and years lived with disability (YLD). YLD for each disease was obtained by multiplying avoided cases by disease-specific average disability weights and durations, sourced from Global Burden of Disease (GBD) 2019 data sources (YLD/prevalence for weights, prevalence/incidence for duration).^27^ Disease durations were applied as constant means by disease, but constrained by remaining life expectancy at advanced ages for consistency.^28^ Total DALYs averted were summed over ages, diseases, and spatial units.

### 2.5 Adjustment of temporality of prospective benefits

To account for delayed health responses following air-pollution reductions, we applied cessation-lag (phase-in) functions that distribute the full epidemiological benefit over time rather than assuming instantaneous reductions. COMEAP Report specified lag structures (e.g., a 20-year lag structure for long-term latency) were applied to reflect biological latency and the delayed reversal of risk for chronic outcomes following exposure reduction.^17^ For each endpoint, avoided deaths and other health outcomes estimated from the concentration-response function were converted into time-specific benefits by multiplying the benefits to a set of lag weights. The detailed methodology of cessation lag adjustment under different assumptions is given in Annex-V. Overall, this approach allows front-loading of outcomes with rapid reversibility (e.g., acute cardio-respiratory mechanisms) while retaining a multi-year effect realizations for conditions with slower progression and long latency (e.g., chronic respiratory disease and cancer). Including cessation lags is consistent with health-economics and burden-of-disease practice. It is critical for correctly interpreting near-term (2030) benefits versus long-term (2050) benefits, as these models reallocate effect timing rather than remove it.

### 2.6 Health Economic Assessment

Disease-specific healthcare costs were sourced from the AMELI French health insurance database (https://data.ameli.fr). Economic benefits of reduced air pollution exposure were evaluated using two approaches. Direct medical costs were based on disease-specific expenditures for outpatient care, hospitalizations, and allowances, given in 2019 Euros (Table S1). Pathology groups included stroke (acute/sequelae), lung cancer, COPD, asthma, type-2 diabetes, and hypertension. For each disease, average annual cost per case was multiplied by avoided cases and average disease duration to estimate total costs prevented. To value mortality and other societal benefits, intangible costs were estimated using the Value of a Statistical Life Year (VSLY): €135k, €152k, and €192k for 2019, 2030, and 2050, respectively per French guidelines.^29^ DALYs were multiplied by VSLY to monetize health benefits. Total benefits equal avoided direct costs plus monetized DALYs.

### 2.7 Uncertainty estimation and sensitivity analysis

A structured sensitivity analysis tested the robustness of health impact estimates to key epidemiological, demographic, and temporal modelling assumptions. First, uncertainty in the concentration–response relationship was addressed by repeating mortality calculations using alternative (more conservative) and locally calibrated relative-risk (RR) functions (Table 2), to check if scenario impacts were sensitive to the risk function, since RRs vary by population, exposure metric, and model. Second, Monte Carlo simulation (1,000 iterations) propagated upper and lower bound uncertainties, sampling RRs from log-normal distributions; outcomes were recomputed each iteration to generate empirical uncertainty intervals (2.5th–97.5th percentiles). Finally, exposure–response temporal dynamics were compared between immediate and delayed-response assumptions.

## 3 Results

### 3.1 Projected PM_2.5_ and NO_2_ concentrations

Across ADEME’s *Transitions 2050* scenarios, projected declines in population-weighted concentrations were broadly similar, with consistently larger relative improvements for NO_2_ than for PM_2.5_ in continental France. For PM_2.5_, the population-weighted mean concentration in S1 decreased from 8·80 µg.m^-3^ in 2019 to 6·70 µg.m^-3^ in 2030 and 5·34 µg.m^-3^ in 2050. These changes correspond to reductions of about 23% by 2030 and 38% by 2050 (Figure 1). For NO_2_, reductions were more pronounced; under S1, population-weighted mean concentrations fell from 11·27 µg.m^-3^ in 2019 to 5·10 µg.m^-3^ in 2030 and 2·51 µg.m^-3^ in 2050. Relative declines reached roughly 45-50% by 2030 and 70-75% by 2050 (Figure 1). Detailed maps of each scenario and their relative changes in concentrations are given in Figure S1 and Figure S2. In particular, national mean NO_2_ concentrations fell below the WHO guideline (10 µg.m^-3^) by 2030 across scenarios, while national mean PM_2.5_ remained slightly above the WHO guideline (5µg.m^-3^) even by 2050. Despite strong improvements in urban centers, gradients persist and concentrations in some cities may remain higher than the national mean.

**Figure 1:**
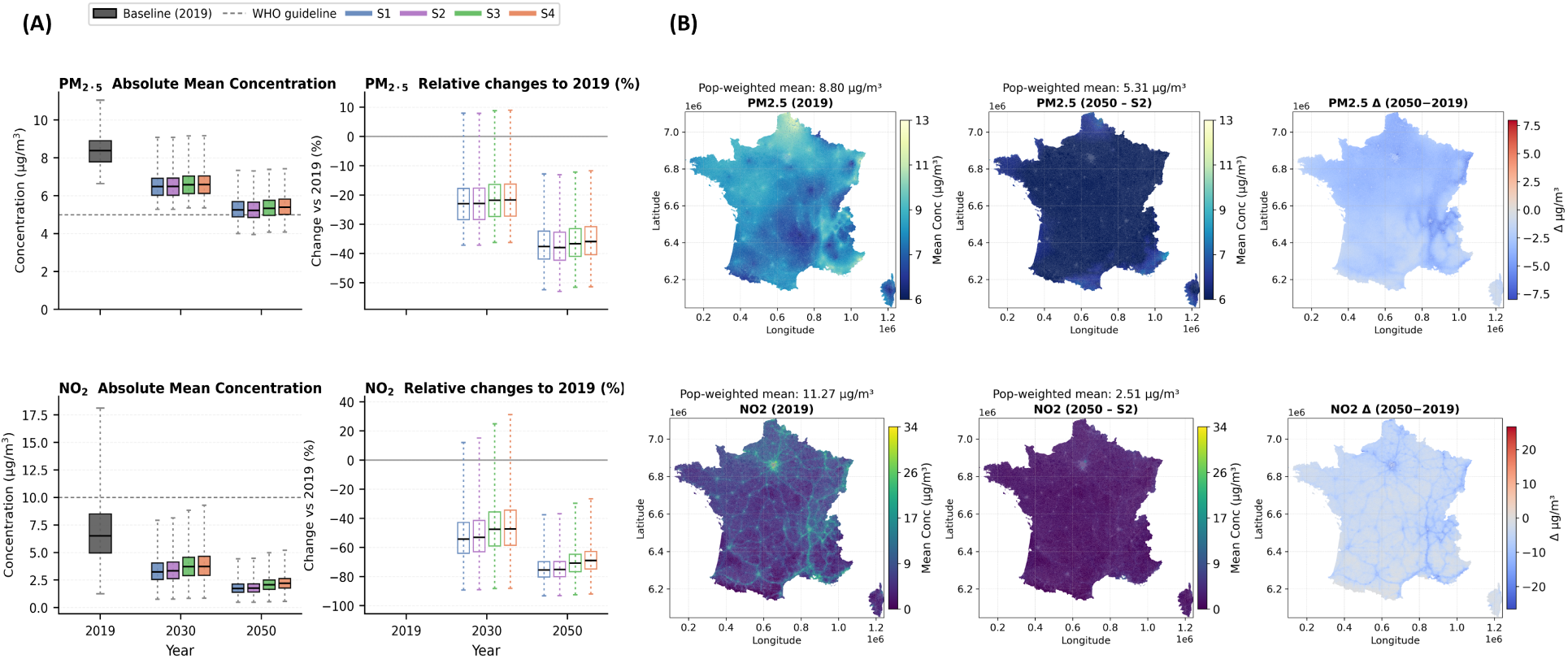
**Spatial distribution of projected annual mean PM**_2.5_ **concentrations (µg.m^-3^) across metropolitan France under ADEME scenarios (S1 to S4) for 2030 and 2050, and the corresponding changes relative to the 2019 baseline (Delta Concentration). (A)** Box plots summarise the distribution of grid-cell mean concentrations across continental France; reflecting Absolute vs. relative change (%) in mean PM_2.5_ and NO_2_ concentrations compared with 2019. **(B)** Spatial distribution of pollutants including 2019 (baseline), scenario-specific (S2) annual mean PM_2.5_ and NO_2_ concentrations in 2050, and delta concentration (2050–2019). *Note: Due to similarities across scenarios, only maps for S2 are shown here in main text*.

### 3.2 Mortality Benefits from Improved Air Quality

Health gains were broadly similar across decarbonisation pathways. We therefore focus on the contrasting S1 and S4 narratives, as S2 and S3 showed comparable intermediate results Figure 2 and Table S2. To contextualise the projected gains, we first estimated the preventable burden under two counterfactual scenarios. Achieving the WHO air quality guidelines in 2019 would have prevented an estimated 22,000 (13,600–28,200) PM_2.5_-attributable and 12,000 (7,800–17,600) NO_2_-attributable premature deaths. Applying a more stringent theoretical minimum-risk counterfactual of 3 *µ*g/m^3^ and 1 *µ*g/m^3^ increased these estimates to 32,400 (20,000–41,200) for PM_2.5_ and 31,900 (19,600–43,600) for NO_2_, respectively.

**Figure 2:**
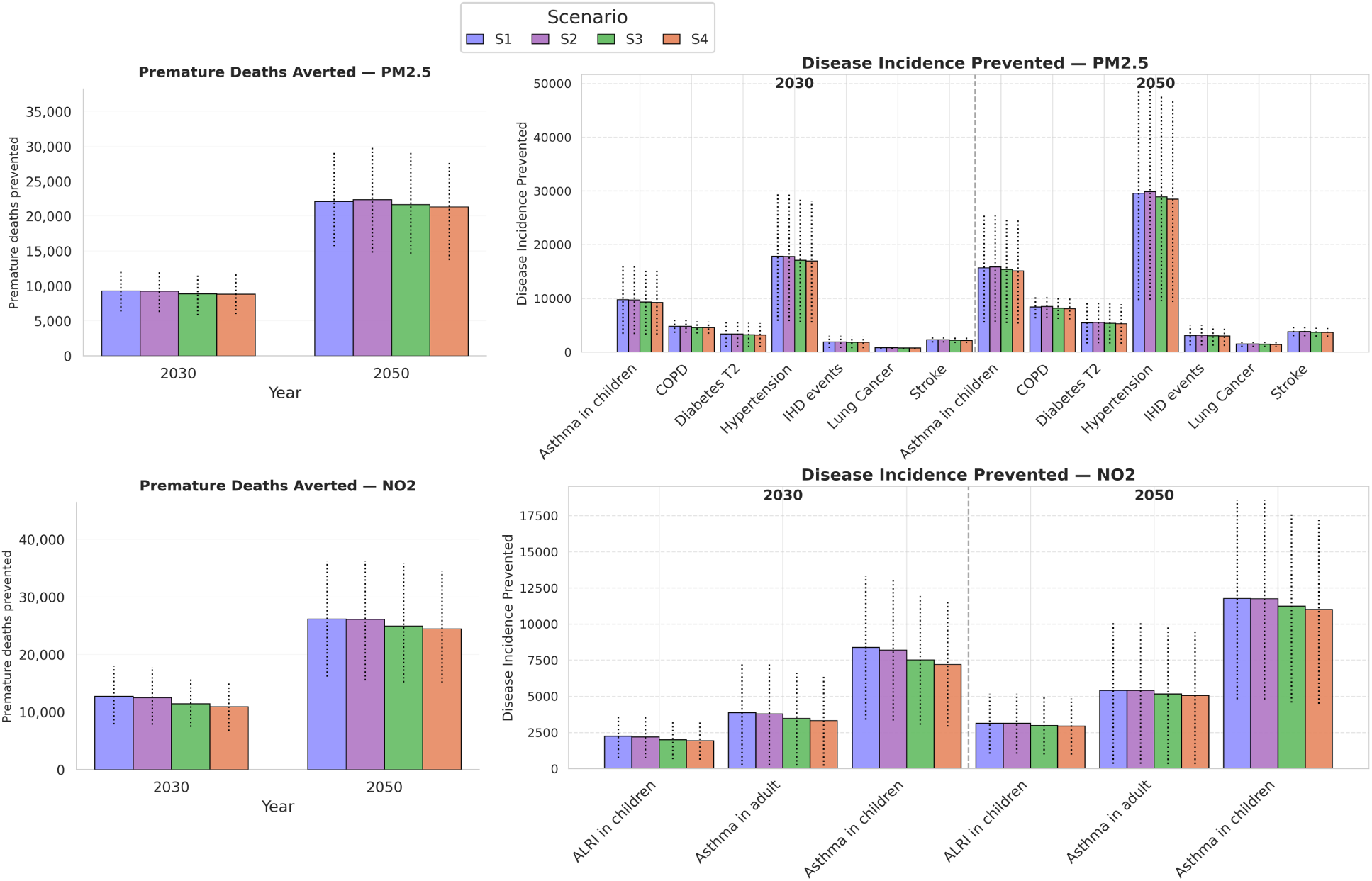
Projected premature mortality and disease cases prevented among adults (*>*30 years) and children in continental France under ADEME Transitions 2050 scenarios. Bar plots show (left) premature deaths averted and (right) incident disease cases prevented for major morbidity outcomes due to reductions in PM_2.5_ (top row) and NO_2_ (bottom row) in 2030 and 2050, relative to a 2019 baseline. Results are shown for all four net-zero scenarios (S1–S4), with dotted whiskers indicating 95% uncertainty intervals.

Health benefits were estimated under two assumptions: an *immediate-response* model and a *cessation-lag* model, which spreads mortality reductions over several years after exposure changes. The cessation-lag results are presented as the primary analysis throughout. Avoided premature deaths for PM_2.5_ ranged from 9,300 (6,100–12,400) in S1 to 8,800 (5,800–11,600) in S4 in 2030, increasing to 22,100 (14,200–29,300) in S1 and 21,300 (13,900–28,300) in S4 by 2050. For NO_2_, avoided deaths ranged from 12,800 (7,700–17,900) in S1 to 11,000 (6,900–15,200) in S4 in 2030, rising to 26,200 (16,900–36,300) in S1 and 24,500 (15,200–33,600) in S4 by 2050 (Figure 2; Table S2). Relative to the 2019 French baseline mortality Table S1, these gains represent 1.6% (S1) to 1.5% (S4) for PM_2.5_ and 2.1% (S1) to 1.8% (S4) for NO_2_ in 2030, rising to 3.7% (S1) to 3.6% (S4) and 4.4% (S1) to 4.1% (S4), respectively, by 2050. Using the WHO guideline counterfactual, the 2050 scenarios lead to the mortality benefits equivalent to nearly all PM_2.5_-related health gains (about 96%) and more than all NO_2_-related gains (about 204%).

Averted YLL were strongly age-dependent, peaking in the oldest groups and rising substantially from 2030 to 2050 (Figure S3; Figure S4). Additionally, urban vs rural gradient is shown in Table S3 and Table S4.

### 3.3 Morbidity Benefits: Avoided Illness and Disease Burden

Reductions in PM_2.5_ and NO_2_ resulted in substantial morbidity co-benefits, with only modest scenario differences. Under the scenario S1, total avoided morbidity cases across all endpoints reached approximately 55,200 by 2030 (PM_2.5_: 40,700; NO_2_: 14,500), rising substantially to 87,800 by 2050 (PM_2.5_: 67,500; NO_2_: 20,300). Under the S4 scenario, corresponding totals were modestly lower at 51,100 in 2030 (PM_2.5_: 38,700; NO_2_: 12,500) and 84,000 by 2050 (PM_2.5_: 65,000; NO_2_: 19,000). For PM_2.5_, hypertension contributed most at both time points (S1: 29,600 vs S4: 28,500 in 2050), followed by childhood asthma (S1: 15,700 vs S4: 15,100 in 2050), COPD (S1: 8,400 vs S4: 8,100 in 2050), type-2 diabetes (S1: 5,500 vs S4: 5,300 in 2050), and smaller reductions in stroke, ischaemic heart disease, and lung cancer (Figure 2). For NO_2_, impacts were limited to respiratory outcomes, mostly childhood asthma (8,400 vs 7,200 in 2030; 11,800 vs 11,000 in 2050), with smaller contributions from adult asthma and acute lower respiratory infections in children (Figure 2).

Decomposition of DALYs averted showed mortality (YLL) dominated over morbidity (YLD) for both pollutants. For PM_2.5_, YLL comprised about 75% of total DALYs averted (3:1 YLL:YLD), while for NO_2_, YLL comprised 95%, with YLD negligible (Figure 3). Most gains occurred in older adults, especially ages 60–79 (Figure S3–Figure S4). Total DALYs averted increased from 142,000–150,000 in 2030 to 225,000–235,000 in 2050 for PM_2.5_, and from 136,000–160,000 to 202,000–215,000 for NO_2_ in the same period.

**Figure 3:**
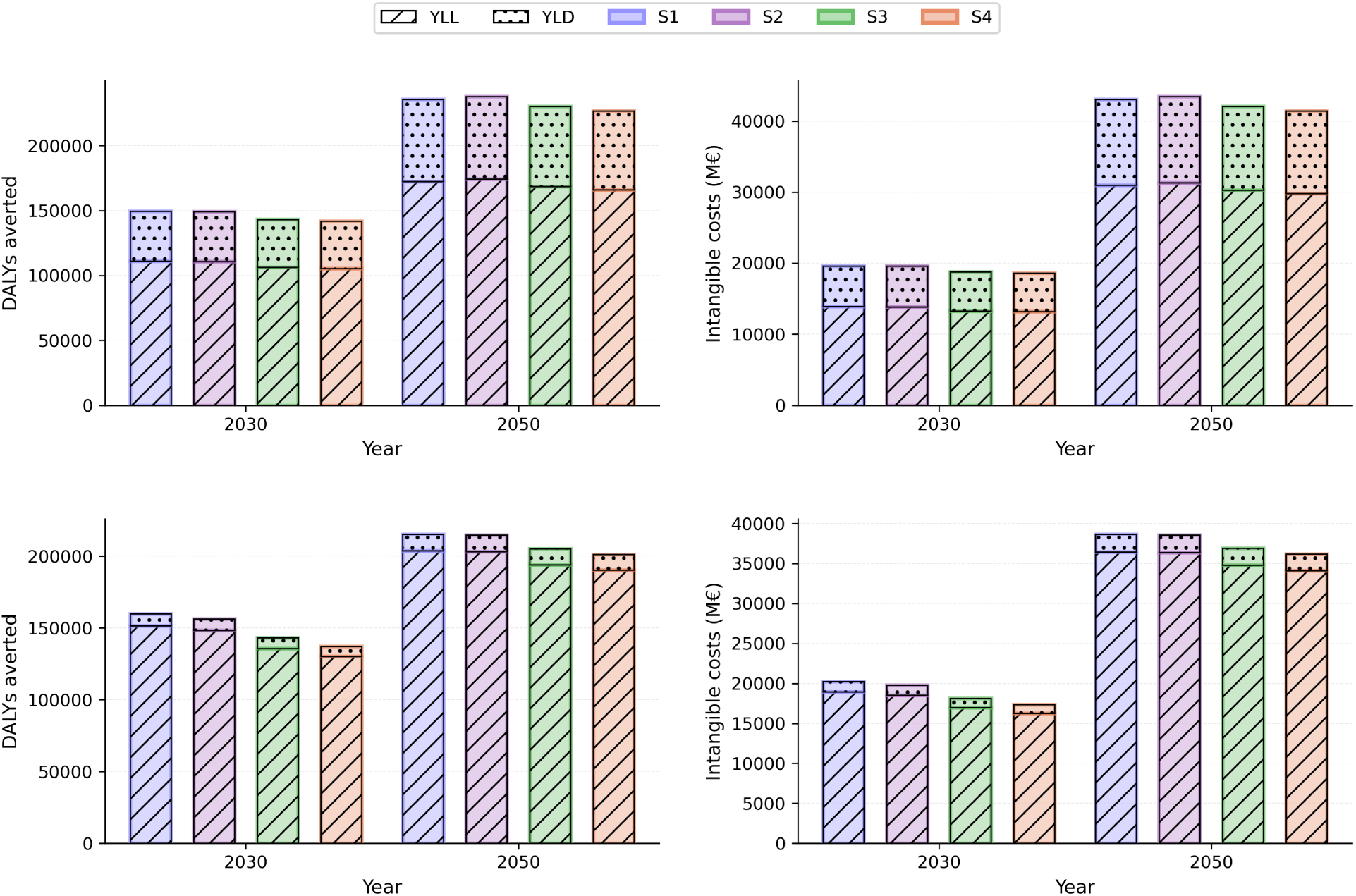
**Projected morbidity benefits associated with reductions in PM**_2.5_ **and NO**_2_ **across continental France under ADEME scenarios (S1 to S4) for 2030 and 2050, relative to the 2019 baseline.** Results report disability-adjusted life years (DALYs) averted (with proportions of mortality vs morbidity), and associated intangible costs prevented (in €M), assuming delayed health impacts. Dotted lines represent uncertainty intervals.

### 3.4 Monetization of the Health Cost

Estimated annual health-care and welfare savings increased substantially with sustained exposure reductions, even under delayed policy impacts (Table 3). For both pollutants together, avoided direct medical costs across all morbidity endpoints were approximately €650 million in 2030 under S1 and €610 million under S4, rising to €1,060 million and €1,020 million respectively by 2050. Avoided intangible costs (morbidity only, YLD-based) were approximately €7,000 million in 2030 under S1 and €6,500 million under S4, rising to €14,300 million and €13,700 million respectively by 2050. For NO_2_, the pattern was similar but at a smaller scale, with direct medical cost savings negligible (ALRI costs not estimated). When mortality-related intangible costs (YLL) are included alongside morbidity, total avoided intangible costs reached approximately €43,000 million (S1) and €41,500 million (S4) for PM_2.5_ alone in 2050, and €38,700 million (S1) and €36,200 million (S4) for NO_2_ in 2050 (Table 3). Hypertension, type-2 diabetes, and stroke accounted for the largest shares of avoided direct medical costs attributable to PM_2.5_ reductions, followed by childhood asthma, whereas for NO_2_, childhood asthma dominated the morbidity cost burden (Figure 3). Had reductions commenced in 2019, cumulative avoided intangible costs accrued up to 2050 would have reached approximately €820–860 billion for PM_2.5_ and €760–850 billion for NO_2_ across scenarios. Cumulative costs were calculated as the sum of annual avoided costs from 2019 to 2050, with annual health benefits evolving according to the cessation-lag structure.

**Table 3:** Projected reductions in health burden and associated avoided costs in 2050 under ADEME S1 and S4 scenarios (delayed impact assumption) in continental France.

| Morbi-mortality event | Scenario S1 |  |  | Scenario S4 |  |  |
| --- | --- | --- | --- | --- | --- | --- |
|  | Cases/deaths prevented [95% CI] | Medical costs, M€ [95% CI] | Intangible costs, M€ [95% CI] | Cases/deaths prevented [95% CI] | Medical costs, M€ [95% CI] | Intangible costs, M€ [95% CI] |
| **PM<sub>2.5</sub>** |  |  |  |  |  |  |
| All-cause mortality | 22,125<br>[15,719–29,317] | – | 30,974<br>[22,004–41,038] | 21,313<br>[13,721–27,639] | – | 29,836<br>[19,206–38,688] |
| Hypertension | 29,551<br>[9,672–48,749] | 286 [94–472] | 6,687<br>[2,170–11,193] | 28,463<br>[9,309–46,987] | 276 [90–455] | 6,441<br>[2,088–10,788] |
| Lung cancer | 1,489 [939–2,031] | 49 [31–67] | 78 [47–112] | 1,434 [905–1,956] | 47 [30–64] | 76 [45–108] |
| Asthma in children | 15,701<br>[5,517–25,440] | 86 [30–139] | 2,014 [672–3,478] | 15,135<br>[5,312–24,555] | 83 [29–135] | 1,941 [647–3,357] |
| COPD | 8,381 [6,290–10,457] | 66 [50–83] | 665 [483–857] | 8,073 [6,057–10,076] | 64 [48–80] | 640 [465–826] |
| Stroke | 3,800 [2,937–4,657] | 149 [115–183] | 1,018 [707–1,369] | 3,659 [2,828–4,486] | 144 [111–176] | 981 [681–1,319] |
| IHD events | 3,100 [1,220–4,943] | 46 [18–74] | 120 [43–210] | 2,986 [1,174–4,762] | 44 [17–71] | 116 [42–203] |
| Type 2 diabetes | 5,470 [1,609–9,276] | 300 [88–508] | 1,475 [413–2,678] | 5,266 [1,548–8,935] | 289 [85–490] | 1,421 [398–2,579] |
| **Total** | – | **982 [426–1,526]** | **43,031**<br>**[26,539–60,936]** | – | **946 [410–1,470]** | **41,450**<br>**[23,572–57,867]** |
| **NO<sub>2</sub>** |  |  |  |  |  |  |
| All-cause mortality | 26,161<br>[16,154–36,191] | – | 36,451<br>[22,505–50,420] | 24,481<br>[15,094–34,487] | – | 34,106<br>[21,026–48,040] |
| Asthma in children | 11,778<br>[4,799–18,608] | 65 [26–102] | 1,511 [582–2,549] | 11,014<br>[4,481–17,428] | 60 [25–95] | 1,413 [543–2,387] |
| Asthma in adults | 5,419 [342–10,215] | 14 [1–26] | 695 [43–1,372] | 5,070 [319–9,579] | 13 [1–24] | 650 [40–1,287] |
| ALRI in children | 3,145 [1,033–5,187] | – | – | 2,940 [964–4,856] | – | – |
| **Total** | – | **78 [27–128]** | **38,657**<br>**[23,129–54,342]** | – | **73 [25–120]** | **36,170**<br>**[21,610–51,715]** |

*Note:* Both scenarios apply cessation-lag adjustments to disease-specific incidence rates, projected to 2050 in continental France and valuation for S2 and S3 scenarios are given in Figure S5. Intangible costs are calculated based on a value of a statistical life year (VSLY) of €135,000 for 2019, €152,000 for 2030, and €192,000 for 2050, with all values expressed in constant Euros 2019. Intangible costs for morbidity endpoints represent the combined YLL and YLD components; intangible costs for all-cause mortality represent YLL only. Medical cost estimates are reported in 2019 Euros without inflation or discounting, representing a conservative estimation approach. ALRI: acute lower respiratory infection; COPD: chronic obstructive pulmonary disease; IHD: ischaemic heart disease; YLD: years lived with disability; YLL: years of life lost.

### 3.5 Uncertainty and sensitivity analysis

Figure 4 summarizes the sensitivity analysis of health benefits from policy-driven PM_2.5_ and NO_2_ reductions. Scenario ranking remained consistent across all variants and pollutants, confirming robustness. Alternative RR assumptions shifted estimates by roughly −50% to +55% vs. the main analysis, while stable-demography assumptions reduced estimates by 15–45%, depending on pollutant and horizon. For internal validation, alternative RRs were also applied to 2016–2019 data using WHO counterfactual thresholds Table S5.

**Figure 4:**
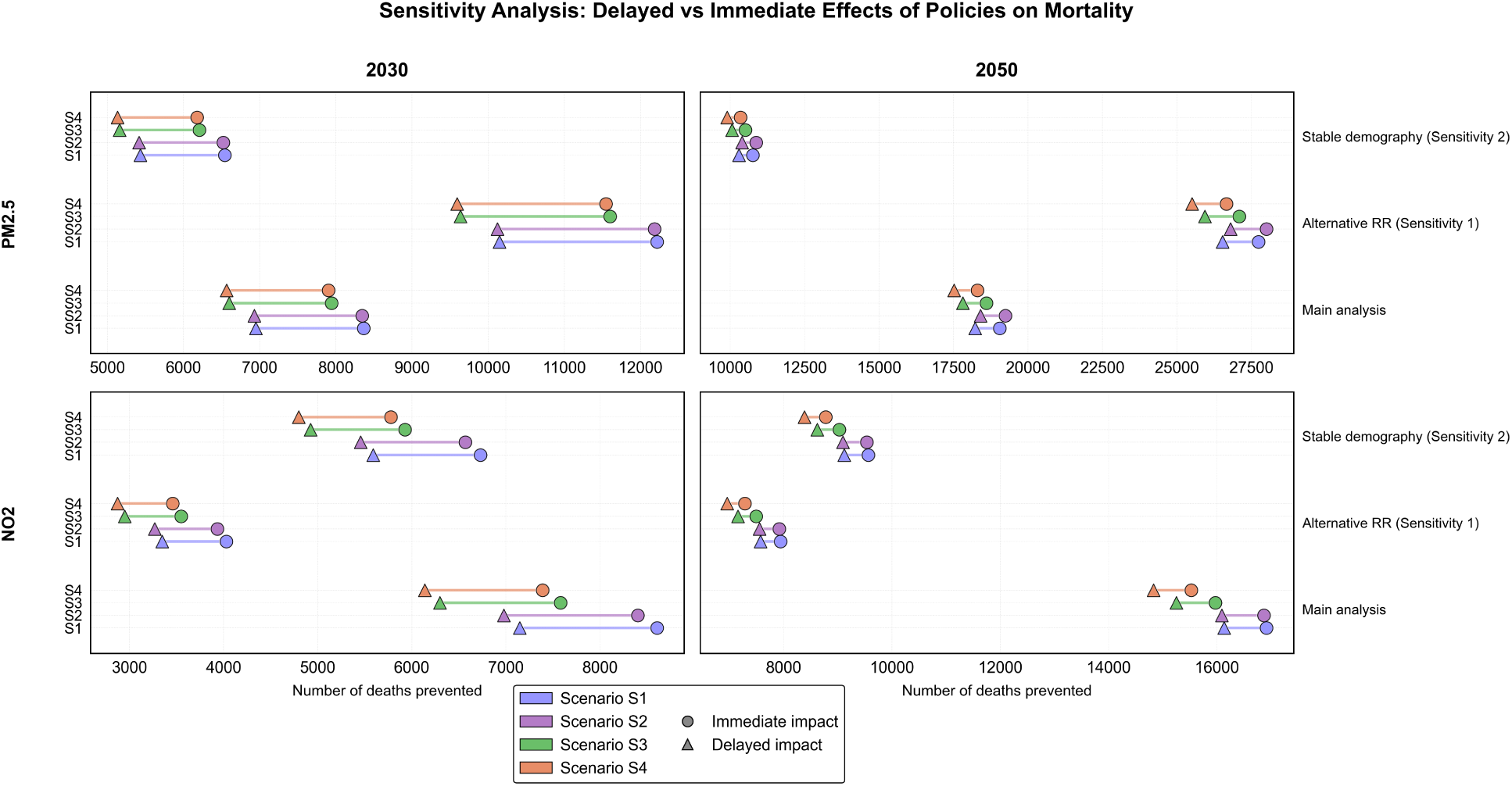
**Sensitivity analysis of projected health and economic benefits from policy-driven reductions in PM**_2.5_ **(top panel) and NO**_2_ **(bottom panel).** Results are shown for the main analysis and two sensitivity variants: (Sensitivity 1) alternative/localised concentration–response (relative-risk, RR) functions and (Sensitivity 2) a stable-demography assumption. For each variant, estimates are compared under assumptions of immediate versus delayed (cessation-lag) realisation of impacts.

Lagged-response estimates were systematically lower than immediate-response estimates in 2030, reflecting biological latency in health benefit accrual. For PM_2.5_, main-analysis premature deaths averted declined from 13,000–14,500 (immediate) to 9,000–10,000 (lagged), a reduction of roughly 30%. For NO_2_, the decline was approximately 20–25%, from 11,000–13,000 (immediate) to 9,000–10,500 (lagged). By 2050, immediate and lagged estimates converged, with differences narrowing to 5–8% for PM_2.5_ and 4–5% for NO_2_, consistent with benefits accruing over time (Figure 4). DALYs showed a similar immediate-lagged gap narrowing from 15–25% in 2030 to 5–8% by 2050 for both pollutants (Figure S6). Time-resolved assessments are in Figure S7, with sensitivity settings detailed in Annex-V.

## 4 Discussion

In this study relying on projected changes in pollutant emissions, fine-scale air pollutant concentrations from a chemistry-transport model, and a health impact assessment, we show that contrasted climate mitigation options may translate into consistent and sizeable mortality and morbidity health co-benefits in continental France. Under the delayed-impact assumption, PM_2.5_ reductions could prevent approximately 8,800–9,300 premature deaths in 2030, rising to 21,300–22,100 by 2050, while NO_2_ reductions could avert 11,000–12,800 deaths in 2030 and 24,500–26,200 by 2050. Although PM_2.5_- and NO_2_-attributable deaths cannot be directly summed because of potential overlap in their health effects, the combined reductions suggest that, under a conservative interpretation, the total mortality benefits by 2050 are on the order of 36,000–48,000 premature deaths averted. Morbidity co-benefits were similarly large, with total avoided cases reaching approximately 55,000–87,000 across scenarios and time horizons. DALY decomposition shows mortality accounts for most averted burden, the increasing YLD contribution by 2050 highlights an accumulating quality-of-life dividend central to health-system planning, yet often absent from scenario appraisals. Economically as well majority of the projected cost-benefits are from the intangible value of avoided premature mortality rather than medical cost savings.

Although the ADEME *Transitions 2050* scenarios were designed as NZ pathways rather than air quality targets, they generate meaningful air pollution co-benefits, while also revealing residual gaps. Under all scenarios, a ∼40% reduction in PM_2.5_ is projected by 2050. Nevertheless, the national mean is projected to remain above the WHO’s guideline of 5 µg.m^-3^ implying that concentrations in several cities and sub-national regions could still substantially exceed this threshold. In contrast, mean NO_2_ concentrations are projected to fall by nearly 70% by 2050, meeting the WHO guideline of 10 µg.m^-3^. However, the log-linear exposure-response relationship^30,31^ assumes linearity near guideline levels Table S2, consistent with the WHO-HRAPIE framework,^18^ meaning even modest reductions at low baselines retain meaningful health relevance. This divergence reflects differences in pollutant sources and formation pathways: NO_2_ is a primary combustion pollutant and responds rapidly to transport and industrial controls, whereas PM_2.5_ arises from both primary emissions and secondary formation from precursor gases (NO*_x_*, SO_2_, VOCs and NH_3_) across transport, buildings, industry, and agriculture, making reductions more diffuse and slower. This is especially relevant as ADEME scenarios do not explicitly explain how agricultural NH_3_) emissions will evolve, representing a notable source of uncertainty. The literature further suggests that the hard-to-abate fraction of PM_2.5_ in Europe is strongly driven by secondary inorganic aerosol formation, in which reactive nitrogen (Nr) notably from NO*_x_* and agricultural NH_3_ plays a central role.^32^ These findings call for jointly designed climate and air quality policies: meeting NO_2_ targets alone is insufficient, closing the remaining PM_2.5_ gap likely requires explicit, high-ambition, NH_3_ inclusive measures alongside continued NO*_x_* reductions.^33^

Despite methodological differences, our projected PM_2.5_ decline is broadly consistent with sustainability-oriented pathways reported for high-income European settings.^34^ Relative to some prospective estimates for the UK, which project smaller proportional declines in population-weighted PM_2.5_ and NO_2_ by 2050, our findings imply greater mid-century potential for PM_2.5_ improvement in France; moreover, explicit accounting of population growth and aging could likely provide increased absolute gains by 2050, but it is a known fact that demographic change could attenuate relative risk reductions.^35^

Economically, the majority of projected benefits (97%) derives from the intangible value of avoided premature mortality rather than direct health-care cost savings, approximately 40:1 for PM_2.5_ alone in 2050. The similar ratios in France were also seen for health co-benefits from physical activities (ratio of 25:1).^28^However, intangible costs carry less political weight than budgetary savings despite representing the largest welfare share; narrowly focusing on healthcare expenditure risks systematic undervaluation of mitigation benefits. Direct medical savings alone are nonetheless economically meaningful. Cumulating annual avoided costs from 2019 to 2050 that is, summing each year’s health gains as exposure falls and disease burden gradually declines, yields savings of approximately €21 billion in direct medical costs over this period. When welfare losses are included, cumulative intangible savings reach approximately €820–860 billion for PM_2.5_ and €760–850 billion for NO_2_ across scenarios. These figures are economically significant for France’s estimated ecological transition financing needs of €110 billion by 2030.^36^ Our findings suggest that air quality improvements alone could generate cumulative direct medical cost savings of nearly €21 billion over 2019-2050-budgetary gains, when intangible gains are also incorporated into cost-benefit appraisals, these gains could offset a considerable share of decarbonisation investment.^37^ Literature further suggests monetised health co-benefits may exceed mitigation costs in the second half of the century (2050-2100),^38^ highlighting that each year of delayed action forfeits welfare gains.

Importantly, national mean estimates could mask substantial regional variation. Background pollution levels, meteorological conditions, demographic structure, and cross-border transport all influence how exposure reductions translate into health gains, leading to spatially heterogeneous outcomes.^39^ City-scale assessments further support this pattern, with preventable mortality burdens ranging from negligible to high depending on local conditions.^40^ As a result, some urban areas may remain above WHO guideline levels despite large absolute reductions, influencing health gains.

Our analyses yield several methodological insights for prospective health impact assessments. The preserved scenario ordering within each pollutant across both time horizons suggests pathway differences reflect underlying exposure trajectories rather than modelling choices. Particularly, latency matters for near-term interpretation as delayed-response specifications systematically influence 2030 benefits relative to immediate-response assumptions, indicating that near-term co-benefits can be overstated if lagged assumptions are ignored. Convergence of immediate and lagged estimates by 2050 (typically within 10%) suggests latency assumptions mainly reallocate benefits over time rather than altering long-run magnitude, indicating importance of sustained implementation.

This study compares two contrasting polices, S1 and S2 emphasizes demand reduction, behavioural change, and cooperation-oriented governance, combining sufficiency measures with regulation and stringent standards to constrain high-emitting activities. While, S3 and S4 relies primarily on technology-led decarbonization, focusing on innovation and efficiency improvements, with less change to consumption patterns. These estimates are nonetheless subject to uncertainties inherent in ADEME’s emission projections, which reflect assumptions about technological adoption, behavioural change, and sectoral transition speeds that may not fully materialise. A further uncertainty concerns the concentration-response functions, held constant across all scenarios. In reality, populations in S1 and S2, which benefit from increased active travel and dietary changes, can arrive in future years with better baseline health, potentially increasing response to air quality improvements; conversely, populations in S3 and S4 can benefit from medical advances that reduce the severity of pollution-related diseases independently of exposure reductions. Capturing such interactions would require integrated health-environment modelling beyond the present study’s scope. Particularly, it is important to note that these findings are specific to air quality co-benefits; health impacts across pathways may be more variable when broader determinants like active travel or diet are considered.

Nevertheless, observed pattern has practical policy implications, articulating near-term air-quality improvements may enhance the political feasibility of sufficiency-oriented pathways by offering tangible public-health returns on shorter timescales than climate outcomes alone.^41^ Policy design should therefore pair early, enforceable actions that rapidly reduce urban combustion-related pollution (especially NO_2_) with longer-term, cross-sector strategies that address the precursor mix driving PM_2.5_. Overall, our results indicate that large, durable air-quality and health co-benefits do not require exclusive reliance on optimistic technological trajectories. A balanced strategy that couples robust regulatory frameworks and demand-side measures with targeted technology deployment can secure early NO_2_ improvements while building the cross-sector foundations needed to sustain PM_2.5_ reductions.

There are several limitations that require consideration. First, while the CHIMERE provides robust estimates of pollutant dynamics and changes, its spatial resolution of the simulation introduces uncertainties, especially at finer resolutions. Although spatial bias correction using INERIS meteorological data helps mitigate discrepancies, residual biases may persist, particularly in urban areas where pollutant sources and meteorology may vary more in future when climate forcing is not included. Second, the HIA applies concentration–response functions uniformly within age strata, assuming homogeneous susceptibility across communes, which may under-represent regional heterogeneity in vulnerability due to differences in health status, deprivation, occupational exposures, housing, and healthcare access. Third, for morbidity, temporal dynamics are less well characterized than for mortality; the implementation assumes constant age-specific incidence rates, possibly oversimplifying future trends and changes (e.g., treatment, vaccination, diagnostics, and behavioural adaptation), potentially affecting baseline rates and speed of risk normalization after exposure reductions. Fourth, demographic ageing and population changes are down-scaled proportionally to IRIS units from national projections. This allows commune-level estimation under consistent demographic assumptions but introduces uncertainty as it does not fully capture spatial variation in migration, ageing, and future urban development. Fifth, DALY estimation for incidence-based YLDs uses average disease durations and disability weights from GBD 2019, without explicitly modelling heterogeneity in age at onset, comorbidity, or disease progression, so YLDs may be biased for subgroups with different durations of disability (e.g., much shorter or longer life expectancy).

Sixth, the economic analysis focuses on direct medical expenditure from AMELI data (e.g., hospitalizations, outpatient care, selected allowances), but excludes wider societal costs such as informal caregiving, additional productivity losses, and other welfare impacts. This limitation is partly addressed by valuing health gains through intangible cost metrics, offering a policy-relevant estimate of societal willingness to pay for risk reduction. Finally, as an ecological assessment, individual-level confounding (socioeconomic status, smoking, occupation, lifestyle) cannot be fully controlled; residual confounding may persist when transferring epidemiologically-derived functions to projected sub-national contexts. A key strength of our study is quantification of economic benefits under both timing assumptions, demonstrating that most gains derive from prevented premature mortality, substantiating mitigation as a public-health investment and guarding against undervaluation in budget-focused policy settings.

## 5 Conclusion

All four net-zero pathways examined here deliver substantial, progressive, and broadly comparable health and economic co-benefits, demonstrating that large gains in population health are a general feature of decarbonisation strategies rather than the exclusive outcome of any single policy design. Our findings further suggest that while immediate policy actions and benefits are appealing, but the full health and economic benefits of air quality improvement will be realized progressively over time, supporting the need for early, decisive, and sustained air quality policies to maximize public health and economic gains. From a decision-making perspective, the return-on-investment from air quality policies and sustained progress towards NZ strategies should be framed in terms of maximizing collective health and economic benefits alongside air quality improvements, since the two are intrinsically linked and could translate into greater health and economic gains.

## Supporting information

Annex

## Data Availability

All data produced in the present study are available upon reasonable request to the authors.

## Contributors

Author initials correspond to those in the author list. AS, AG, AC, and KJ contributed to the conceptualisation of the study. AS conducted the formal analysis and visualisation, and wrote the original draft of the manuscript. AS, AG, ER, and MC, MP, SM and VW contributed to data curation and methodology. Funding was acquired by AG, AC, and KJ. AC and KJ supervised the work. All authors contributed to writing – review and editing of the manuscript, and approved the final version for submission.

## Data sharing

Data and analysis scripts used in this study are available at: https://github.com/Sharma19Ayushi/HIA_airquality_PhiloCTET_ATMO

## Supplementary Information

**Table S1:**
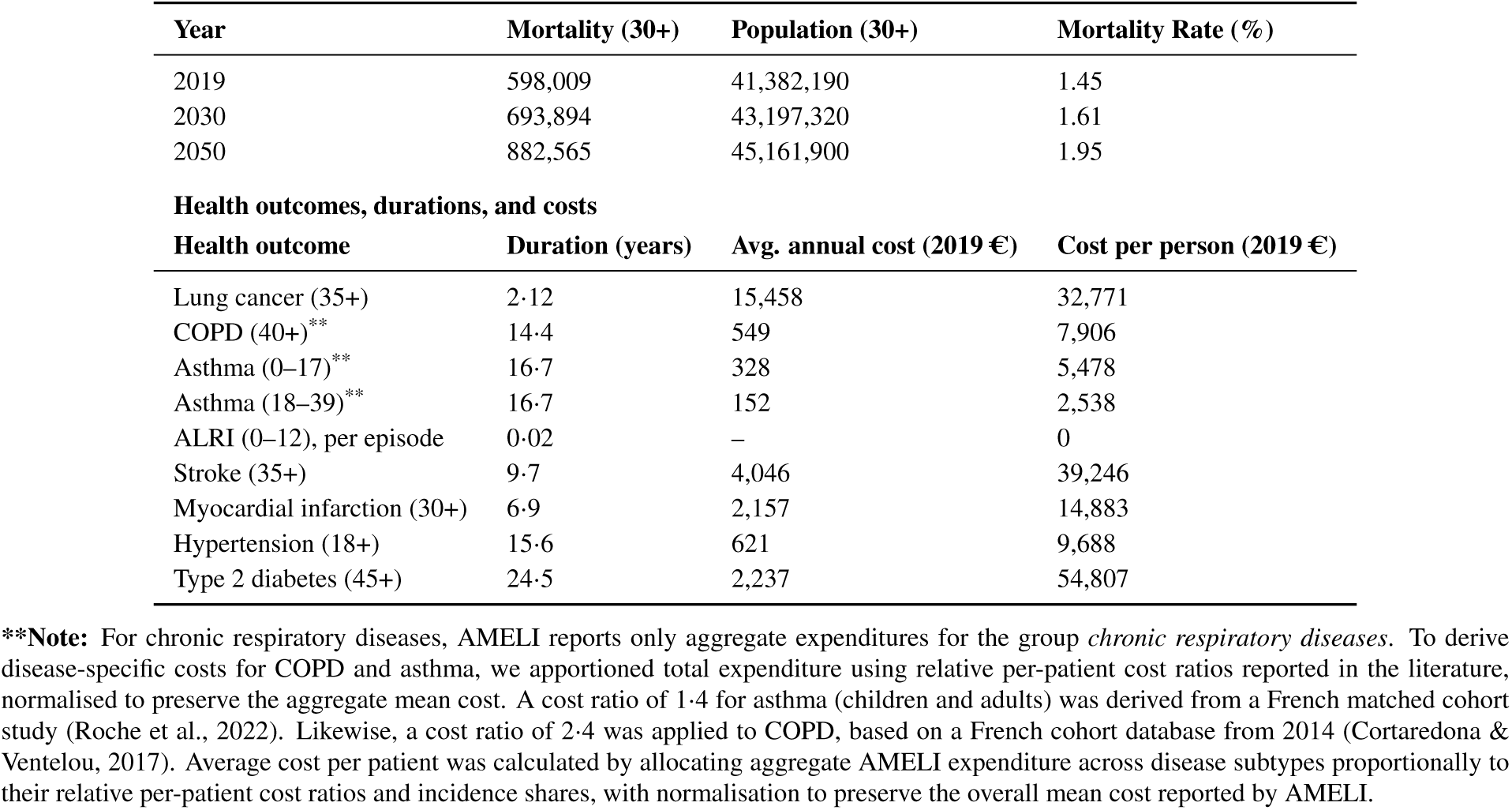
Mortality, population (age 30+), and disease cost summary for France, 2019, 2030, 2050.

**Table S2:** Marginal health benefits of a 0.1 *µ*g/m^3^ reduction in population-weighted commune-level PM_2.5_ and NO_2_ concentrations, using baseline demography (2019) to extract just pollutant effects. Percent change is relative to 2019 baseline.

| Scenario | Pollutant | Year | Mortality prevented |  | Years of life lost prevented |  |
| --- | --- | --- | --- | --- | --- | --- |
|  |  |  | n | (%) | n | (%) |
| Baseline | $\text{PM}_{2.5}$ | 2019 | 667.8 | (0%) | 1203.5 | (0%) |
| Baseline | $\text{NO}_2$ | 2019 | 352.6 | (0%) | 634.8 | (0%) |
| s1 | $\text{PM}_{2.5}$ | 2030 | 682.0 | (+2.1%) | 1637.7 | (+36.1%) |
| s2 | $\text{PM}_{2.5}$ | 2030 | 682.0 | (+2.1%) | 1637.7 | (+36.1%) |
| s3 | $\text{PM}_{2.5}$ | 2030 | 681.5 | (+2.1%) | 1636.4 | (+36.0%) |
| s4 | $\text{PM}_{2.5}$ | 2030 | 681.5 | (+2.1%) | 1636.3 | (+36.0%) |
| s1 | $\text{PM}_{2.5}$ | 2050 | 690.6 | (+3.4%) | 1906.9 | (+58.5%) |
| s2 | $\text{PM}_{2.5}$ | 2050 | 690.8 | (+3.4%) | 1907.6 | (+58.6%) |
| s3 | $\text{PM}_{2.5}$ | 2050 | 690.0 | (+3.3%) | 1905.4 | (+58.4%) |
| s4 | $\text{PM}_{2.5}$ | 2050 | 689.8 | (+3.3%) | 1904.6 | (+58.4%) |
| s1 | $\text{NO}_2$ | 2030 | 364.54 | (+3.4%) | 875.08 | (+37.8%) |
| s2 | $\text{NO}_2$ | 2030 | 364.31 | (+3.3%) | 874.50 | (+37.8%) |
| s3 | $\text{NO}_2$ | 2030 | 363.35 | (+3.0%) | 872.16 | (+37.4%) |
| s4 | $\text{NO}_2$ | 2030 | 362.87 | (+2.9%) | 870.98 | (+37.3%) |
| s1 | $\text{NO}_2$ | 2050 | 369.21 | (+4.7%) | 1019.17 | (+60.6%) |
| s2 | $\text{NO}_2$ | 2050 | 369.18 | (+4.7%) | 1019.08 | (+60.6%) |
| s3 | $\text{NO}_2$ | 2050 | 368.47 | (+4.5%) | 1017.09 | (+60.3%) |
| s4 | $\text{NO}_2$ | 2050 | 368.24 | (+4.4%) | 1016.45 | (+60.2%) |

**Table S3:** Urban–rural gradient in premature deaths prevented attributable to PM_2.5_ reductions across net-zero scenarios, 2030 and 2050.

| Pollutant | Year | Scenario | Area | Premature deaths prevented | Deaths prevented (%) |
| --- | --- | --- | --- | --- | --- |
| PM<sub>2.5</sub> | 2030 | S1 | Urban | 4,476 (2,989–5,814) | 40% |
|  |  | S2 | Urban | 4,464 (2,900–6,048) | 40% |
|  |  | S3 | Urban | 4,274 (2,876–5,663) | 39% |
|  |  | S4 | Urban | 4,242 (2,852–5,657) | 39% |
|  |  | S1 | Peri-urban | 3,740 (2,484–5,020) | 33% |
|  |  | S2 | Peri-urban | 3,731 (2,450–4,982) | 33% |
|  |  | S3 | Peri-urban | 3,585 (2,386–4,767) | 33% |
|  |  | S4 | Peri-urban | 3,557 (2,432–4,718) | 33% |
|  |  | S1 | Rural | 2,977 (2,065–3,978) | 27% |
|  |  | S2 | Rural | 2,969 (2,015–4,007) | 27% |
|  |  | S3 | Rural | 2,851 (1,965–3,840) | 27% |
|  |  | S4 | Rural | 2,829 (1,862–3,794) | 27% |
|  | 2050 | S1 | Urban | 9,262 (6,078–12,336) | 40% |
|  |  | S2 | Urban | 9,357 (6,489–12,336) | 40% |
|  |  | S3 | Urban | 9,054 (6,217–11,842) | 39% |
|  |  | S4 | Urban | 8,913 (6,070–11,853) | 39% |
|  |  | S1 | Peri-urban | 7,715 (5,287–10,196) | 33% |
|  |  | S2 | Peri-urban | 7,797 (5,071–10,441) | 33% |
|  |  | S3 | Peri-urban | 7,547 (4,058–9,970) | 33% |
|  |  | S4 | Peri-urban | 7,436 (5,059–9,955) | 33% |
|  |  | S1 | Rural | 6,159 (4,107–8,185) | 27% |
|  |  | S2 | Rural | 6,224 (4,218–8,166) | 27% |
|  |  | S3 | Rural | 6,022 (4,009–8,106) | 27% |
|  |  | S4 | Rural | 5,932 (4,121–7,943) | 27% |

**Note:** Values are central estimate (95% uncertainty interval). Urban, peri-urban, and rural classifications follow the INSEE–Eurostat typology applied to French communes. Percentage of deaths prevented is relative to the pollutant-attributable mortality baseline within each area type. PM<sub>2.5</sub> counterfactual: 5 µg/m<sup>3</sup>.

**Table S4:**
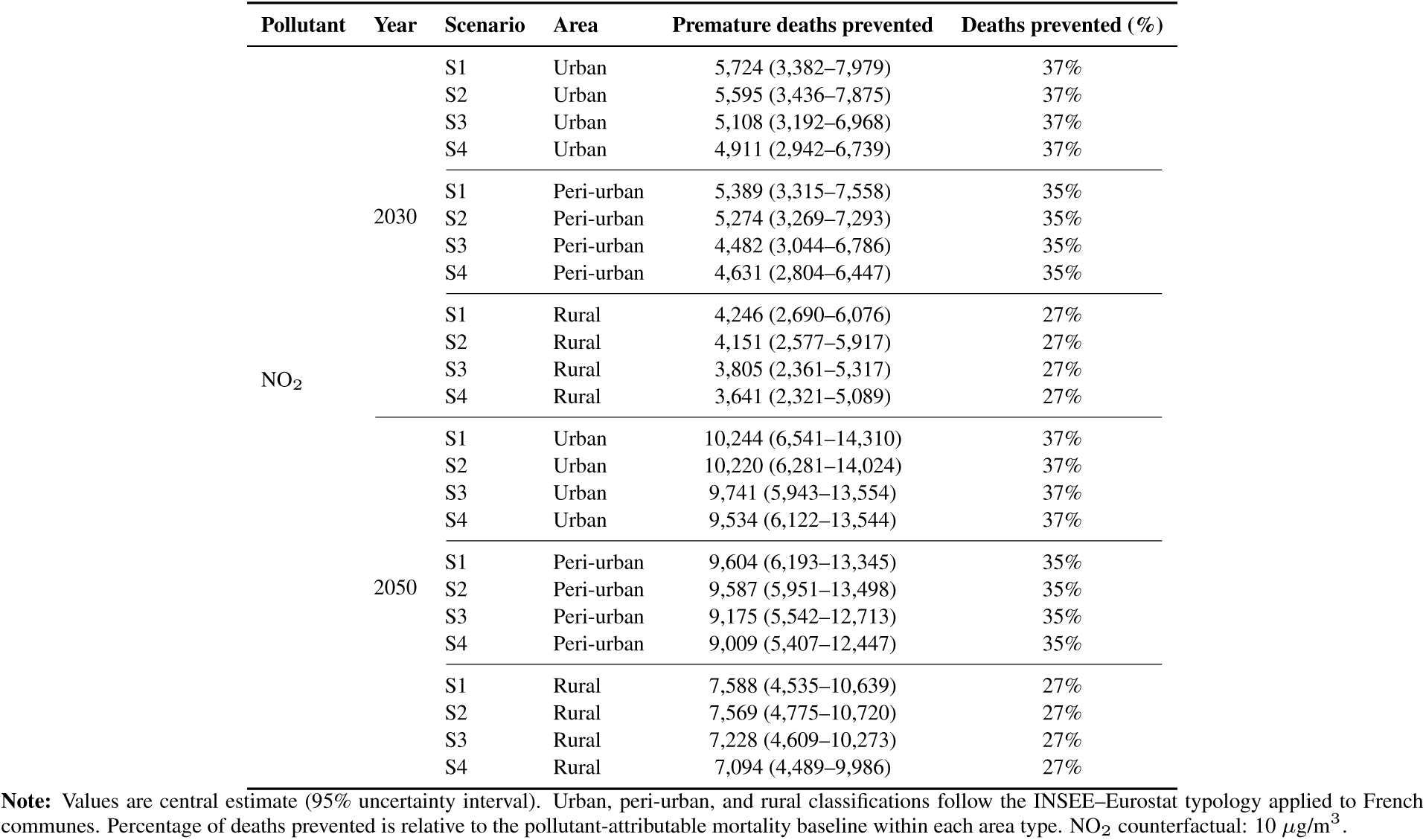
Urban–rural gradient in premature deaths prevented attributable to NO_2_ reductions across net-zero scenarios, 2030 and 2050.

**Table S5:**
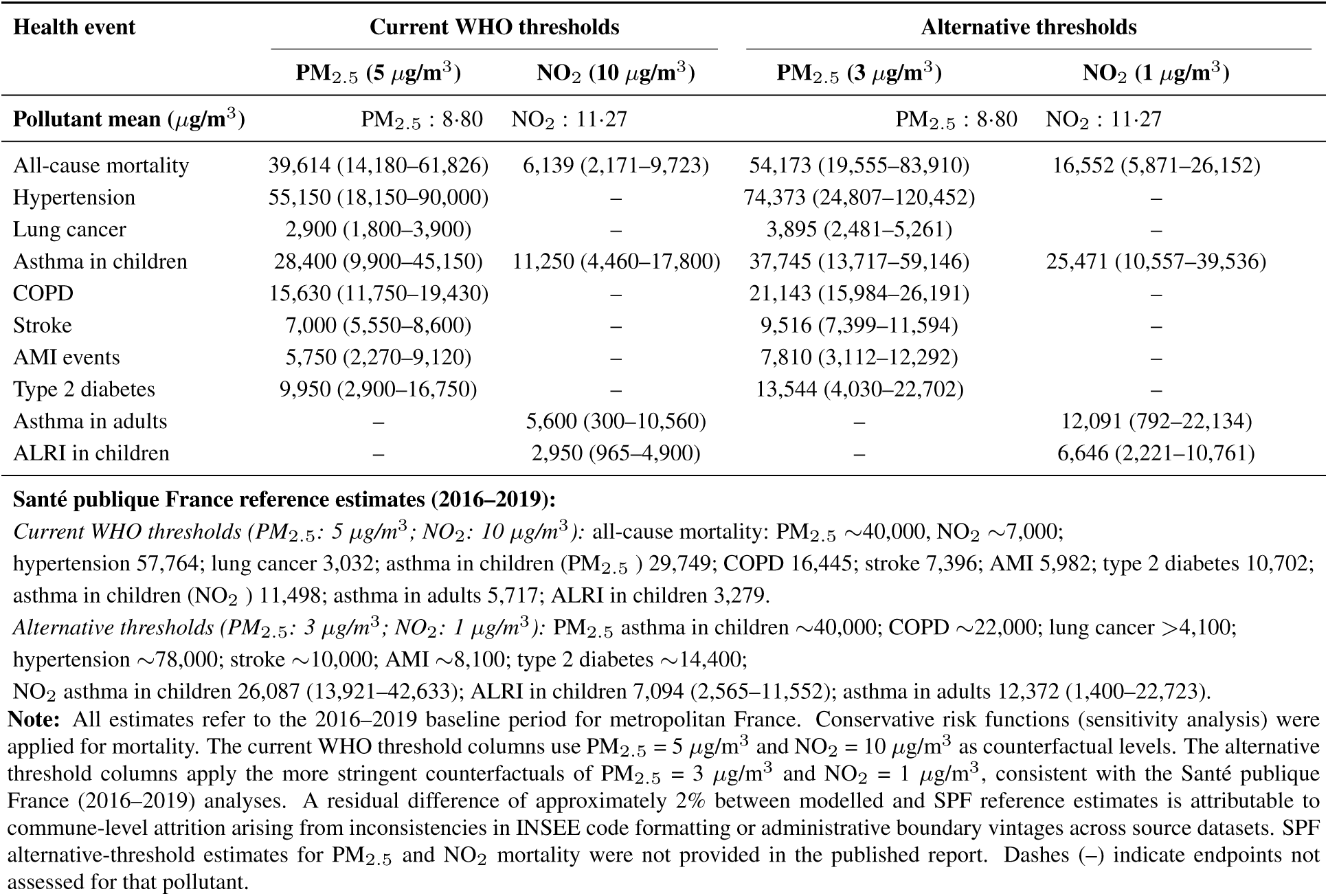
Internal validation–comparison of baseline-year (2016–2019) modelled outcomes with Santé publique France published estimates under WHO and alternative counterfactual concentration thresholds.

**Figure S1:**
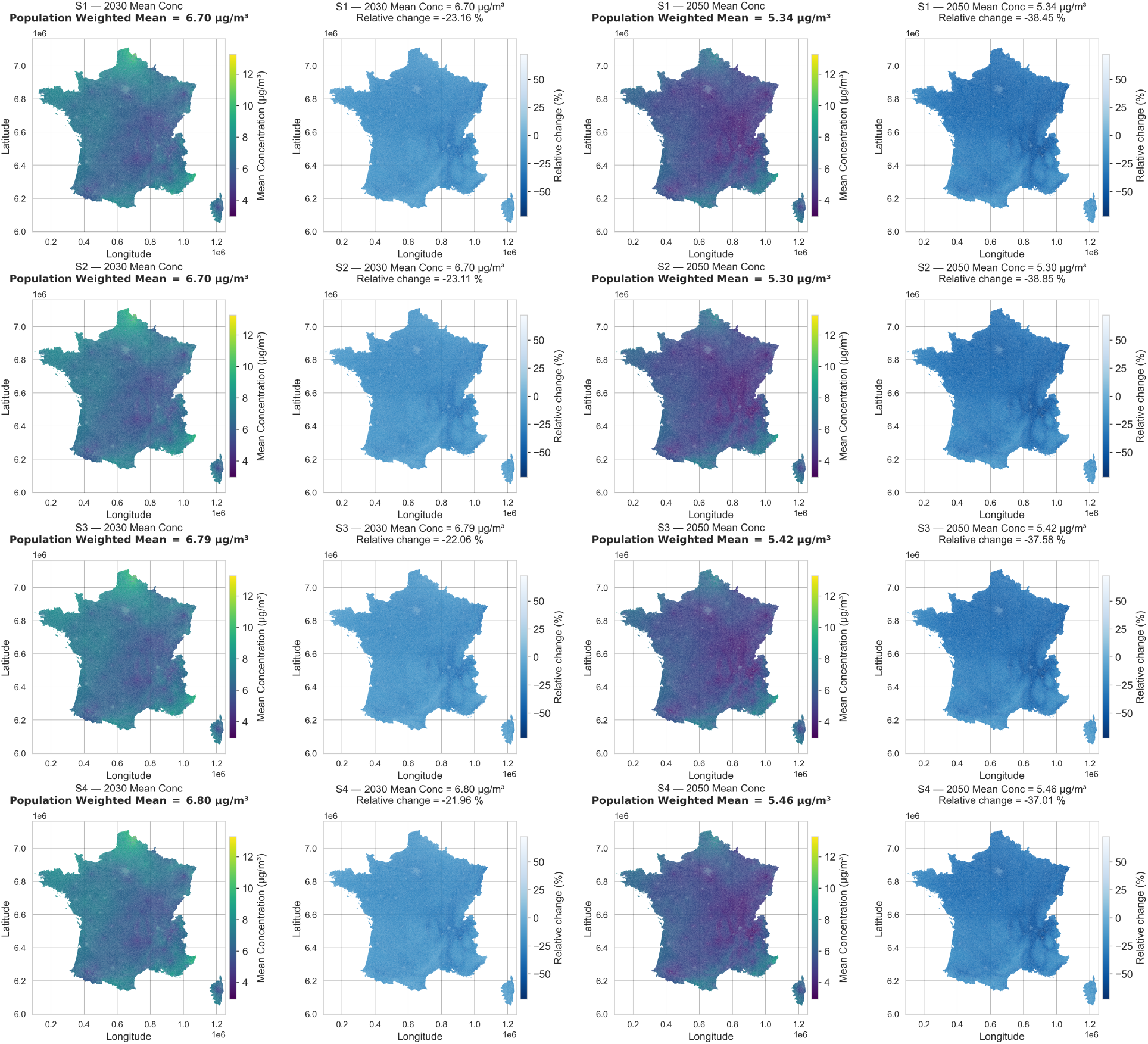
Maps showing the spatial distribution of projected annual mean PM_2.5_ concentrations (*µ*g/m^3^) and relative changes (% variation from 2019 concentration levels) across continental France under ADEME *Transitions 2050* scenarios, for 2030 and 2050.

**Figure S2:**
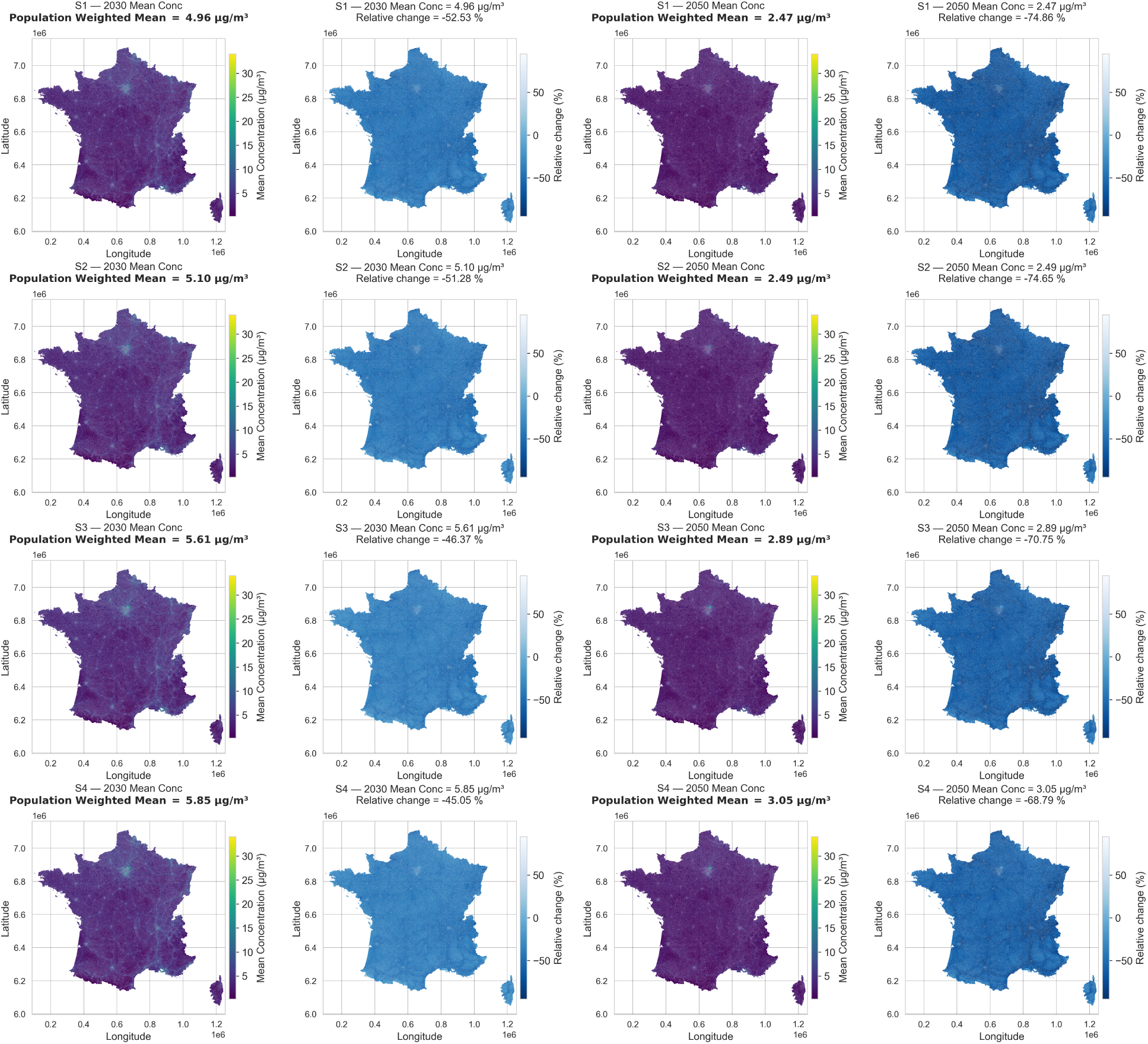
Maps showing the spatial distribution of projected annual mean NO_2_ concentrations (*µ*g/m^3^) and relative changes (% variation from 2019 concentration levels) across continental France under ADEME *Transitions 2050* scenarios, for 2030 and 2050.

**Figure S3:**
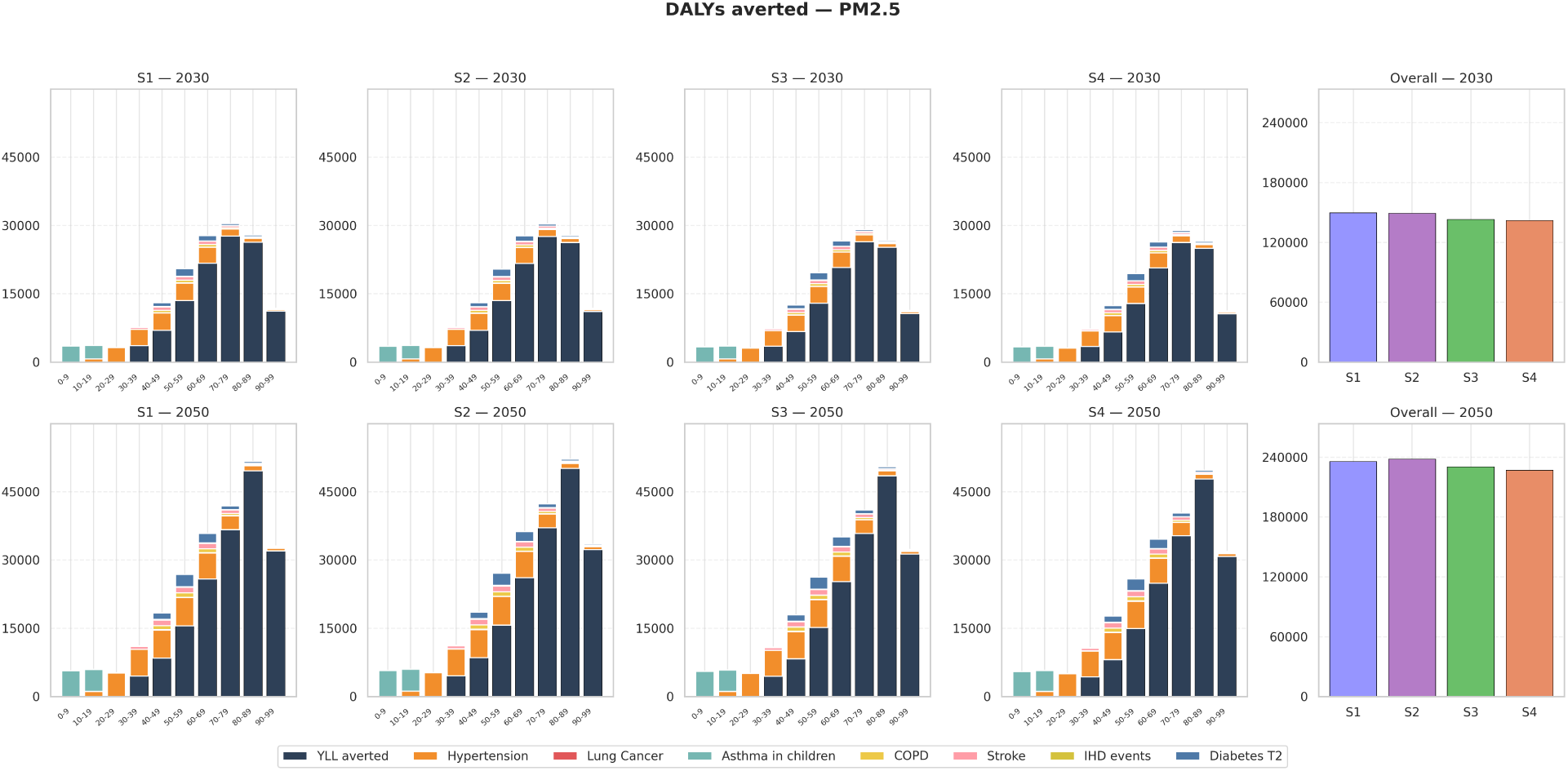
Age-specific disability-adjusted life-years (DALYs) averted, decomposed into years of life lost (YLL) and years lived with disability (YLD), associated with reductions in PM_2.5_ across continental France under ADEME *Transitions 2050* scenarios in 2030 and 2050, relative to the 2019 baseline.

**Figure S4:**
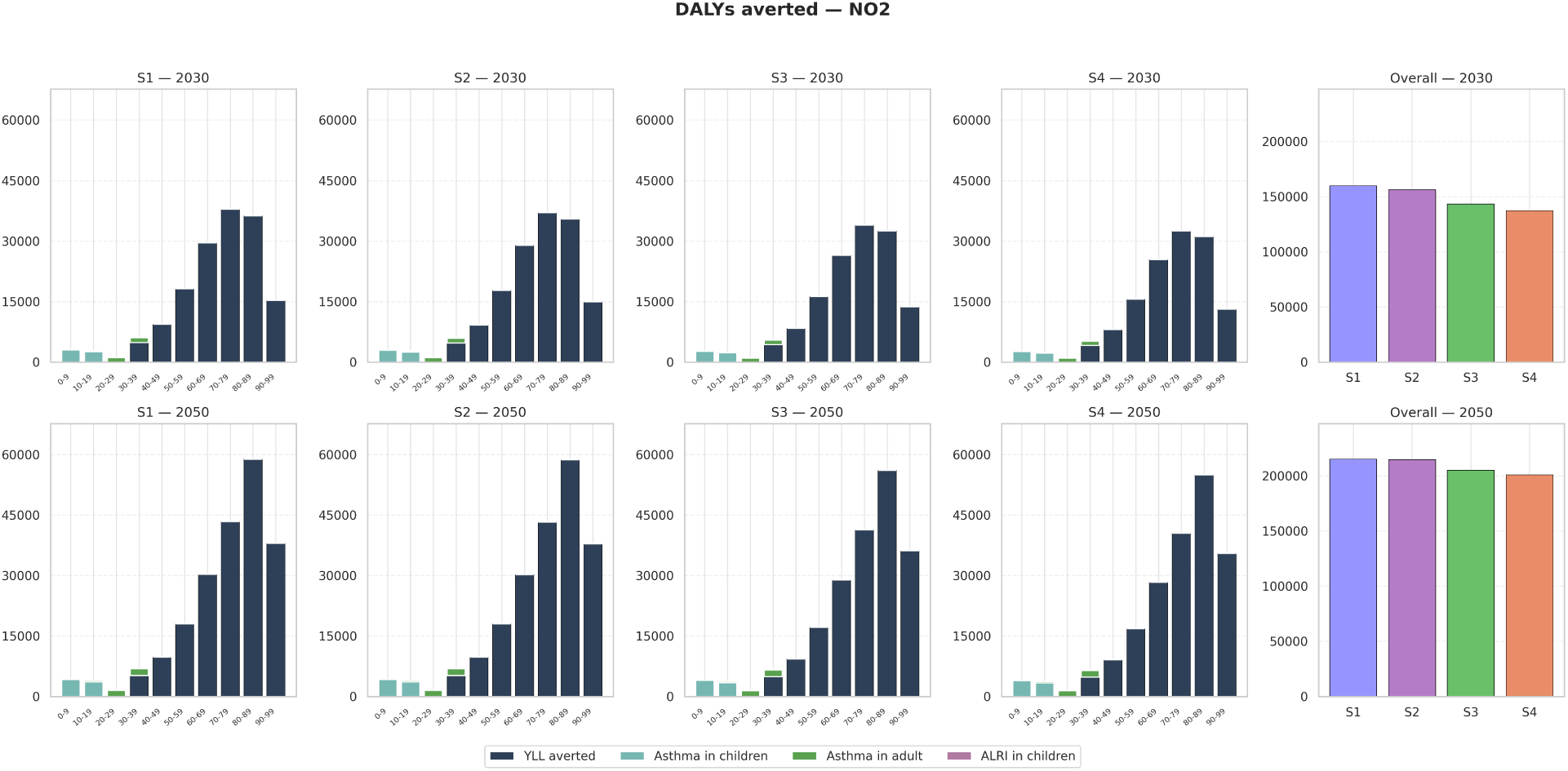
Age-specific disability-adjusted life-years (DALYs) averted, decomposed into years of life lost (YLL) and years lived with disability (YLD), associated with reductions in NO_2_ across continental France under ADEME *Transitions 2050* scenarios in 2030 and 2050, relative to the 2019 baseline.

**Figure S5:**
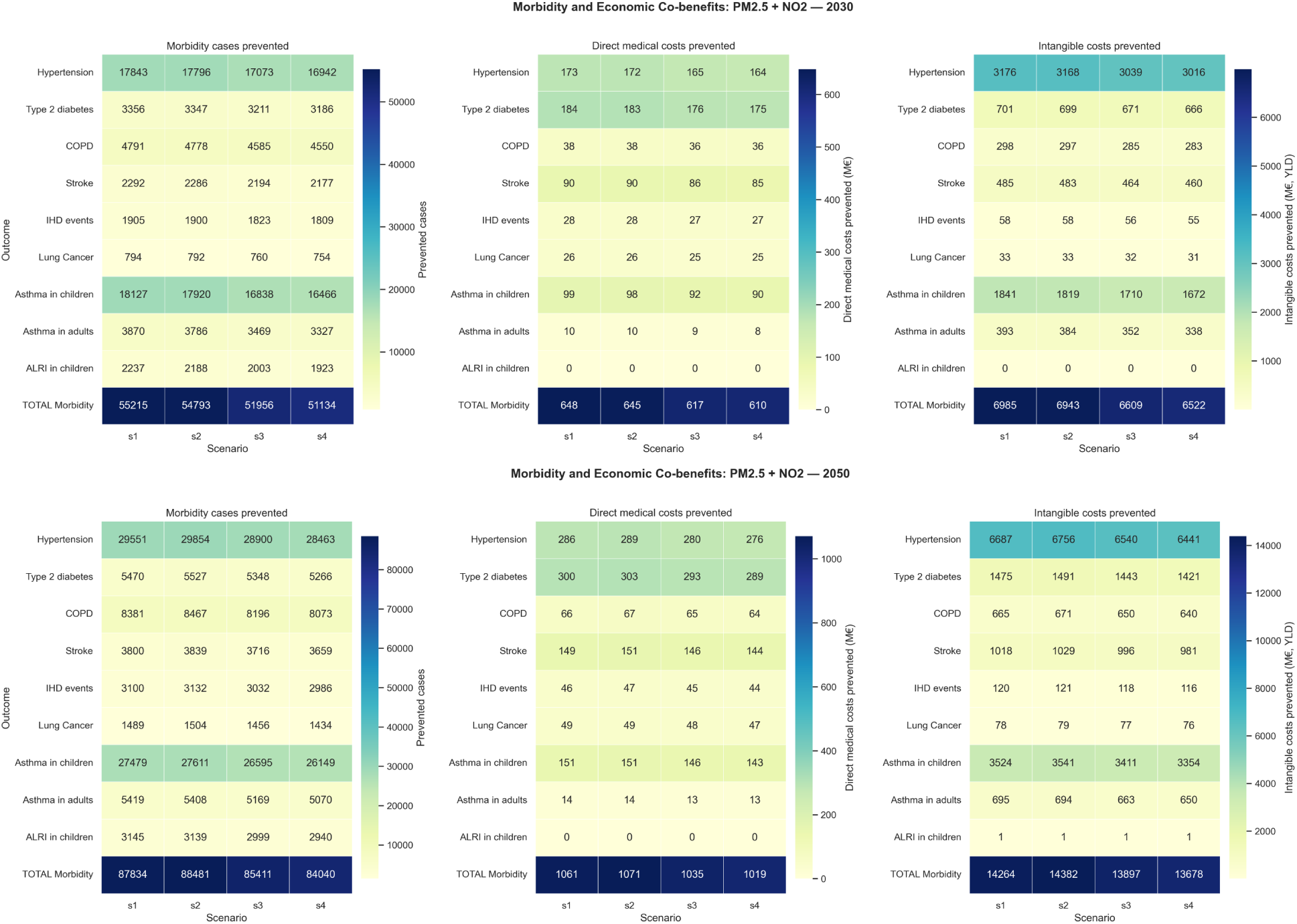
Heatmaps summarising health and economic co-benefits from PM_2.5_ and NO_2_ reductions under ADEME *Transitions 2050* scenarios (S1 to S4) in 2030 (top) and 2050 (bottom), relative to the 2019 baseline. Outcomes include prevented events (cases/deaths), avoided direct medical costs (M€), and avoided intangible costs (M€). Totals shown exclude mortality benefits to avoid double counting across outcome categories.

**Figure S6:**
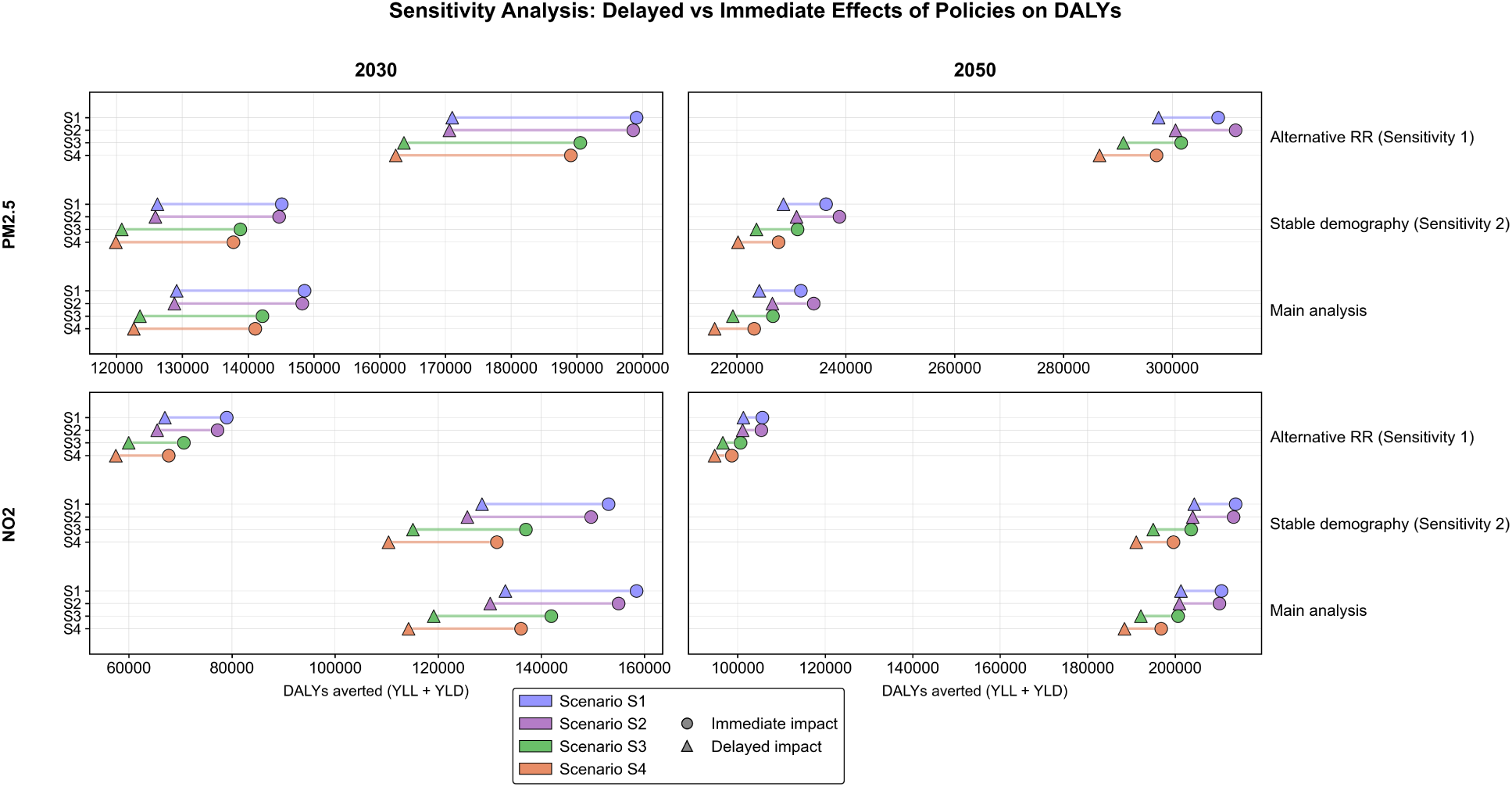
Sensitivity analysis of disability-adjusted life-years (DALYs) averted from policy-driven reductions in PM_2.5_ (top) and NO_2_ (bottom). Results are shown for the main analysis and two sensitivity variants: (Sensitivity 1) alternative/localised concentration–response (relative-risk, RR) functions and (Sensitivity 2) a stable-demography assumption. For each variant, estimates are compared under assumptions of immediate versus delayed (cessation-lag) realisation of impacts.

**Figure S7:**
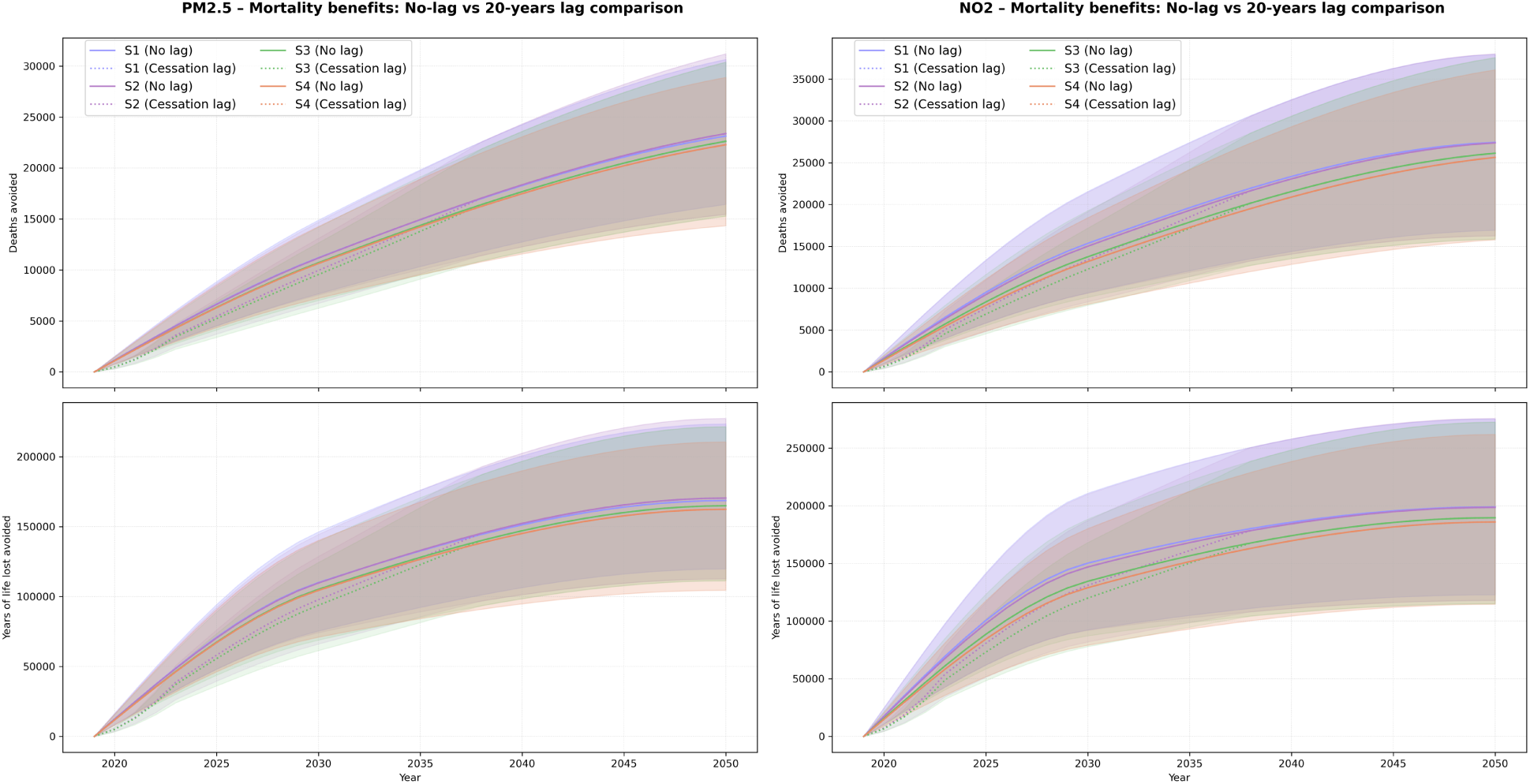
Sensitivity of mortality benefits to lag structure: temporal evolution of cumulative avoided deaths (top) and years of life lost (YLL; bottom) for PM_2.5_ and NO_2_ under ADEME *Transitions 2050* scenarios (S1 to S4). Solid lines indicate no-lag assumptions, whereas dotted lines represent a 20-year cessation lag. Shaded regions denote uncertainty intervals.

