## Supplementary material for "Health and Economic Benefits of Air Quality Improvements in France through Net-Zero Transition Scenarios by 2050": Annex

### ANNEXURE FILE

#### Annexure I: Detailed summary of sectoral transitions in ADEME's prospective scenarios by 2050

| Dimension | S1- Frugal Generation | S2- Regional Cooperation | S3 - Green Technologies | S4 - Restoration Gamble |
|---|---|---|---|---|
|  | -------Sobriety driven scenarios------- |  | -------Technology driven scenarios------- |  |
| **General societal narrative** | Sobriety-driven transformation focused on resource conservation and self-restraint. | Collaborative transition based on shared, responsible consumption. | Technology-driven transformation with advanced living modifications. | Innovation-led restoration relying on engineering solutions and financial investments. |
| **Residential (Buildings)** | Massive rapid renovation; Strong limitation of new builds (conversion of vacant homes and secondary residences into primary residences) | Massive renovation, gradual but profound changes in lifestyles (more developed cohabitation and adjustment of housing size to household size) | Large-scale deconstruction–reconstruction; all housing renovated but in low-performance manner | New construction maintained; Only half of the housing is renovated; Equipment is multiplying, combining technological innovations and energy efficiency |
| **Transport (Mobility)** | Strong reduction in mobility; –33% km/person; 50% trips walking/cycling | Controlled mobility; –17% km/person; ~50% walking/cycling | State-supported mobility infrastructure, +13% km/person; 30% walking/cycling | Very high mobility; +28% km/person; 20% walking/cycling |
| **Food** | Organic food production: 70%; –67% meat consumption; | Organic food production: 50%; –50% meat consumption; | Organic food production: 30%; –30% meat consumption | Meat consumption is almost stable (-10%), compensated by plant proteins |
| **Technology** | Low-tech, repair, reuse; Stable data-center consumption | Massive investment (energy efficiency, renewable energy, and infrastructure); Digital technology serving territorial development; Stable data-center consumption | Competitive decarbonisation tech; Digital tech serving optimization; Data-center energy ×10 vs 2020 | Innovations focussed: Carbon Capture and Storage (CCS); The Internet of Things and artificial intelligence everywhere; Data-center energy ×15 vs 2020 |
| **Governance** | Local decisions, weak international coordination; Demetropolization to medium-sized cities and rural areas | Shared governance; Environmental taxation and redistribution; National decisions and European cooperation; Territorial cooperation | Minimal regulation, state planning, targeted carbon pricing; Metropolization | Centralised energy planning, targeted international cooperation; Small territorial size, urban sprawl, intensive agriculture |
| **Industry** | Local production; ~70% of steel and other materials from recycling | Local markets prioritized; ~80% recycled steel/materials; investment in value than volume | Energy decarbonization; ~60% recycled steel/materials | CCS-based decarbonization; ~45% recycled steel/materials |

#### Annexure II: Methodology for downscaling population and mortality to IRIS spatial unit

The process of downscaling population and mortality data from the national and departmental levels to the IRIS level is built upon the projections and data provided by the National Institute of Statistics and Economic Studies (INSEE). INSEE conducts population projections every five years using the component method, which involves projecting changes in fertility, mortality, and net migration independently. The current analysis is based on INSEE's 2021 study, "*Population Projections 2021-2070 for France*," focusing on the central hypothesis, further details and data can be accessed at: https://www.insee.fr

1. Fertility Rate: The central hypothesis assumes that the fertility rate stabilizes at 1.80 children per woman from 2022 onwards, with the average age of motherhood increasing to 33 years by 2052.
2. Mortality Rate: Death rates by sex and age are assumed to decline at the same rate observed between 2010-2019 (excluding COVID-19 impacts). Generations born between 1941 and 1955 are expected to see continued stagnation in mortality rates. These assumptions predict that life expectancies at birth will reach 90.0 years for women and 87.5 years for men by 2070.
3. Net Migration: The net migration balance is assumed to remain constant at +70,000 individuals per year until 2070.

**Methodology for Obtaining Geographic Data from INSEE Data**

The downscaling process integrates data from multiple INSEE and National Institute of Geographic and Forestry Information (IGN) datasets to derive population and mortality estimates at the granular IRIS level.

**Dataset utilized:**

1. Population Projection Study Published in 2021:
   - Population distribution by age at the national level.
   - Mortality rates categorized by age and sex at the national level.
   - Yearly population projections (2019–2070) for departments, categorized by 5-year age groups.
2. Infra-Municipal Census (IRIS Level) for 2019: detailing the population census data for each IRIS for 2019.
3. Geographical Coordinates of IRIS: A dataset produced by IGN/INSEE that provides spatial geometries and areas for IRIS regions.

**Steps of Data Integration and Processing:** The methodology for downscaling population and mortality data to the IRIS level involved several structured steps to integrate data from multiple spatial and temporal scales. As a first step, IRIS-level shapefiles were merged with demographic data to associate 2019 population records with individual IRIS regions. The integrated dataset included information such as geographic boundaries, population counts, and areas at the levels of departments, regions, communes, and IRIS units.

National-level population data by age and gender, with the five-year age cohorts were disaggregated to obtain granular population distributions for single-year ages. Special algorithms and adjustments were implemented to ensure accurate representation of later age groups, such as 95–99 and 100+. This provided precise proportions of the population within each age bracket, enabling the subsequent steps of the analysis. At the departmental level, population projections for the years 2020 to 2050 were categorized into 5-year age groups and aggregated to ensure consistency across years and regions. The national and departmental population trends were used as the basis for further disaggregation to the IRIS level. To obtain IRIS-level population projections for each year, proportional scaling between departmental and national population growth trends was employed. The IRIS-level population totals for 2019 served as a baseline for the projections, which were adjusted for yearly changes in departmental-level population and national-level age distributions. This method allowed the estimation of age-specific IRIS-level population distributions between 2019 and 2050. Mortality rates at the IRIS level were estimated by aligning national mortality rates by age and sex with the IRIS-level population data. Proportional relationships between national, departmental, and IRIS-level data were used to calculate IRIS-level mortality projections for each age group, integrating changes in population size and national mortality trends.

The choice to downscale population and mortality data to the IRIS level, as opposed to the commune level, is primarily driven by the need for finer spatial granularity. Although the final health impact assessment (HIA) results are reported at the commune level, population and mortality data were first downscaled to the IRIS level to ensure methodological robustness, reduce spatial bias, and improve exposure attribution. Communes in France often encompass substantial internal heterogeneity in population density, age structure, land use, and environmental exposure. Conducting all demographic and health calculations directly at the commune level implicitly assumes spatial homogeneity within communes, which is rarely valid particularly in urban and peri-urban contexts. This assumption can lead to exposure misclassification and biased health impact estimates when environmental risk factors vary at finer spatial scales.

By performing the demographic and mortality allocation at the IRIS level, the analysis captures within-commune variability in population distribution and age structure before aggregation. Environmental exposures are thus linked to populations at the most detailed granular scale, ensuring that health impacts are attributed where people actually live rather than being uniformly distributed across entire communes. From a methodological standpoint, this approach aligns with standard practices in spatial epidemiology and environmental HIA, where calculations are performed at the finest available resolution and results are aggregated to policy-relevant units. Reporting at the commune level ensures interpretability and consistency with administrative decision-making, while IRIS-level processing ensures scientific accuracy. Finally, IRIS-level estimates can be re-aggregated to alternative spatial scales (communes, departments) or reused in sensitivity analyses without recalculating the entire demographic framework. The commune-level HIA results therefore reflect a deliberate analytical choice rather than a data limitation.

**Mathematical equations:**

**1. Disaggregation of Population for 2019**

$$P_{2019}^{\mathrm{IRIS}}(\mathrm{n}) = P_{\mathrm{IRIS,tot}}^{2019} \times {P_{DEP,([5\mathrm{i},5(\mathrm{i}+1)])}} \Big/ {P_{DEP,\mathrm{tot}}^{2019}} \times F^{2019}(\mathrm{n})$$

**2. Downscaling of Future Population Projections**

$$P_{\mathrm{IRIS}}^{\alpha}(\mathrm{n}) = P_{\mathrm{IRIS,tot}}^{2019} \times {P_{DEP,([5\mathrm{i},5(\mathrm{i}+1)])}^{\propto}} \Big/ {P_{DEP,([5\mathrm{i},5(\mathrm{i}+1)])}^{2019}} \times \frac{F^{\alpha}(\mathrm{n})}{F^{2019(\mathrm{n})}}$$

**3. Mortality Estimation at the IRIS Level**

$$M_{\mathrm{IRIS}}^{\alpha}(\mathrm{n}) = P_{\mathrm{NAT}}^{\alpha} \times \frac{P_{\mathrm{IRIS}}^{\alpha}(\mathrm{n})}{P_{\mathrm{NAT}}^{\alpha}(\mathrm{n})}$$

*where,*

***α****: Year (2020 ≤ α ≤ 2050) ;* ***n=5<sub></sub>*** 

**P** *: Population*

**$F^{\alpha}$ (n)***: National fraction of population at age n within the corresponding 5-year age group in year α*

***IRIS*** *: Small-area spatial unit ;* ***DEP****: Département level ;* ***NAT****: National level*

**Key assumptions of the methodology:**

1. Homogeneous Age Distribution: It is assumed that within a given 5-year age group for a specific department, the proportion of each specific age is approximately homogeneous at the national level.
2. 2019 Departmental to IRIS Homogeneity: Within a department in 2019, the age distribution is assumed to be homogeneous across all IRIS regions in that department.
3. Proportional Growth: Population growth at the IRIS level in any given year is assumed to be proportional to the growth observed at the departmental level in that year.
4. Mortality Homogeneity: The mortality rates by age and sex at the national level are assumed to be applicable across

These assumptions simplify the methodology by reducing the complexity, without such simplifications, the calculations would require granular data on age-specific population, mortality, and pollutant exposure for each level (IRIS, department, etc.), which is often unavailable.

***Figure AI.1*** illustrates a comparison of population distributions by age for 2019, 2030, and 2050, derived from IRIS-level population downscaling. These distributions correspond to national population projections from INSEE, demonstrating the robustness and reliability of the methodological approach. A key trend observed is the significant increase in the population aged 70 and above by 2050, highlighting the effect of improved life expectancy and healthcare services of the population. Concurrently, the population of younger age groups (0–20 years) shows a slight decline across the decades, reflecting the stabilization of fertility rates at approximately 1.80 children per woman post-2022, consistent with INSEE projections. The population within age groups 25–60 remains relatively stable over time, aligned with assumptions of gradual aging within the working population and the stability of net migration trends.***Figure AI.2*** presents gender-stratified population pyramids for the same periods, capturing demographic shifts over the decades. The pyramids indicate a widening peak for older age groups, underscoring the increasing longevity predicted by INSEE. Women consistently outnumber men in age groups above 70, consistent with the higher predicted life expectancy of women by 2050. While, gender differences are evident in older populations, the sex ratio is relatively balanced overall, confirming the limited necessity for gender-specific analysis in this study.

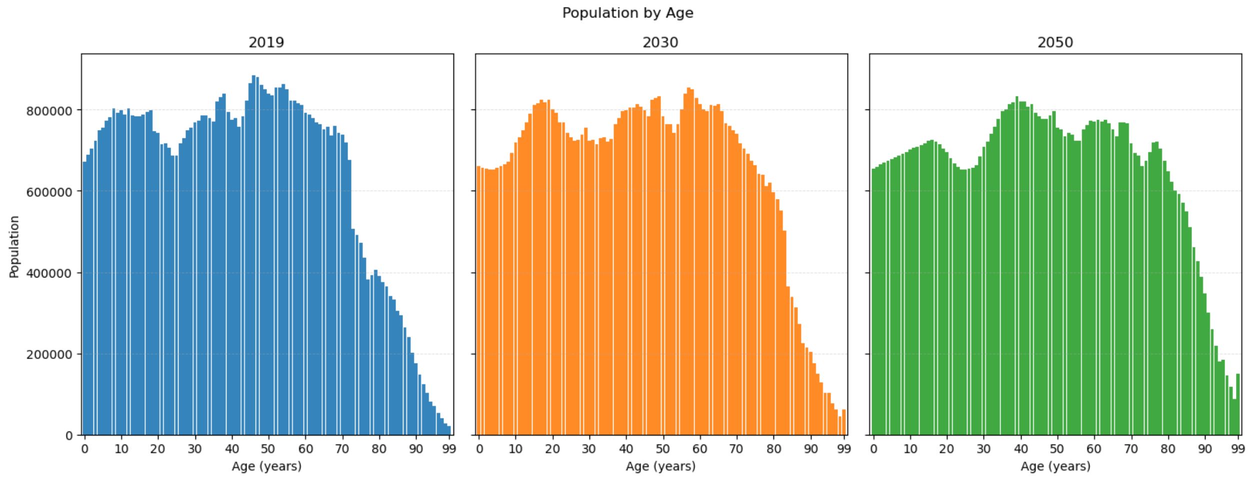


**Figure AI.1- Changes in age-structure of population in metropolitan France for 2019, 2030 and 2050**

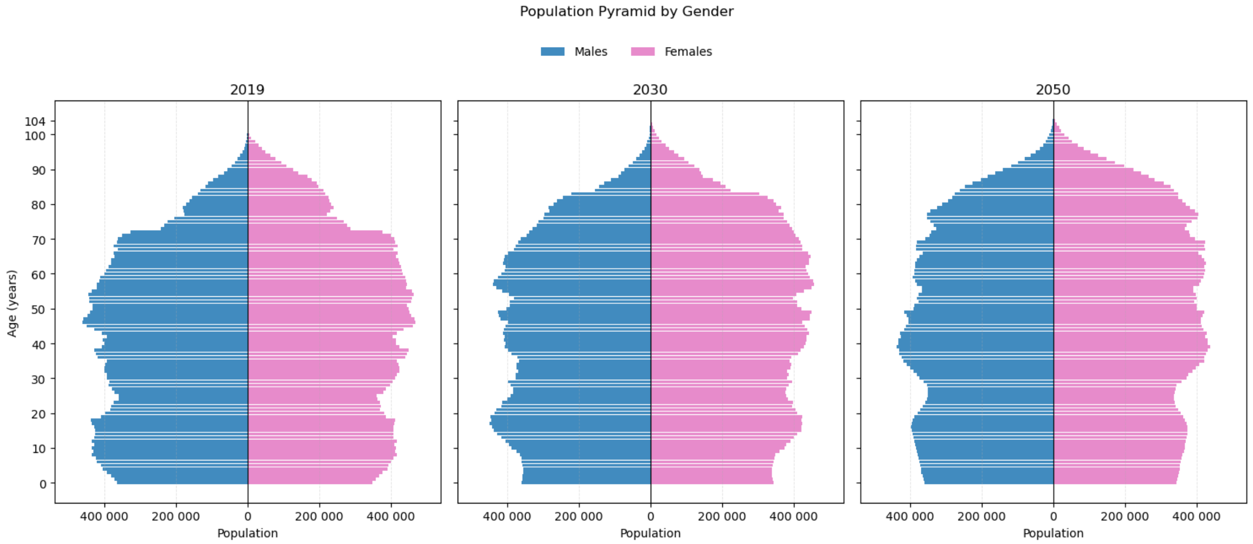


**Figure AI.2 – Gender-stratified population pyramids of metropolitan France for 2019, 2030 and 2050**

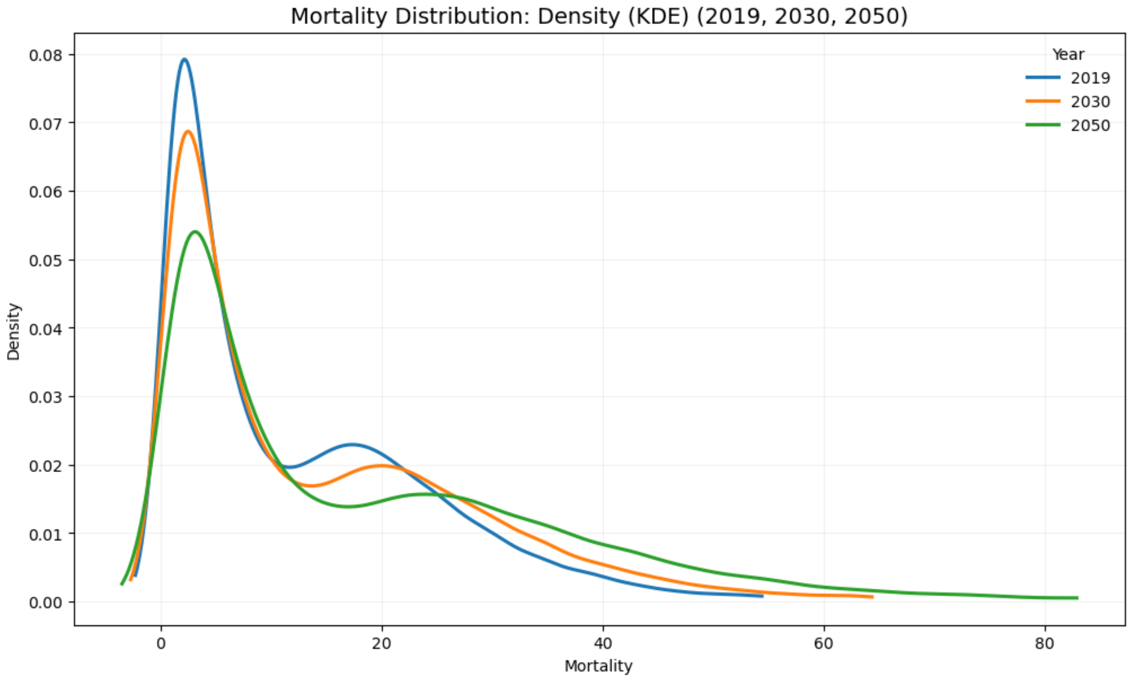


**Figure AI.3- Distribution of mortality over the years 2019, 2030, and 2050 in metropolitan France**

***Figure AI.3*** shows the evolution of mortality distributions for France across the years 2019, 2030, and 2050 using Kernel Density Estimation (KDE). The density curves depict a shift in peak mortality values toward higher ranges by 2050, reflecting the influence of population aging trends. In 2019, the distribution is characterized by a pronounced peak at lower mortality values, indicating a younger population with fewer deaths occurring in older age groups. By 2030, this peak becomes broader and shifts slightly higher, signaling the progression of aging within the population and an increase in age-specific mortality rates. By 2050, the density curve flattens and extends further into higher mortality values, capturing the continued effects of population aging and increased longevity. While fewer deaths occur at lower values, the frequency of higher mortality levels rises, due to an expanding elderly demographic with improvements in life expectancy. This trend aligns with national demographic projections that predict older populations remaining the dominant contributors to mortality rates by mid-century. Together, all these figures accentuate the importance of adopting dynamic population models for future projections. Relying on static demographic assumptions risks underestimating the effects of aging and changes in age-related mortality trends. Dynamic models that incorporate evolving demographics provide more practical, robust insights, especially when planning resource allocations for healthcare systems and air quality management in aging societies.

#### Annexure III – Sensitivity tests and assumptions for distribution of population across age

The methodology described in ***Annexure I*** adopts heterogeneous spatial attribution at the IRIS-level population disaggregation while maintaining homogeneous assumptions for age distribution to simplify certain computations. The ***figure AIII.1*** presents a sensitivity analysis conducted on baseline (2019) air quality data at the commune level, evaluating the potential Years of Life Gained (YLG) due to reductions in $PM_{2.5}$ and $NO_2$ concentrations. The analysis applies the World Health Organization (WHO) guidelines of 5 $\mu g/m^3$ for $PM_{2.5}$ and 10 $\mu g/m^3$ for $NO_2$ as thresholds to assess the mortality impact of improved air quality. Two approaches are compared:

1. **Homogeneous Age-Based Distribution:**
    - This method assumes uniform age distribution across communes.
    - Predicted YLG due to reductions in $PM_{2.5}$ ranges between 217,742 and 953,651, with a central estimate of 609,752 years.
    - For $NO_2$ reductions, YLG spans from 34,145 to 154,643, with a central estimate of 97,102 years.
2. **Spatial Attribution of Age Proportions:**
    - This method accounts for spatial variability in age proportions across communes for a more location-specific analysis.
    - YLG estimates for $PM_{2.5}$ range from 218,002 to 954,789, yielding a similar central value of 610,479 years compared to the first method.
    - For $NO_2$, the projected YLG varies from 34,186 to 154,827, producing a central estimate of 97,218 years.

The graphical subpanels highlight age-specific trends in YLG considering the reduction of pollutants. The panel (a) shows a smooth decline in YLG with increasing age, consistent across conditions, whereas the panel (b) emphasizes the spatial variation and its nuanced influence, particularly at older ages.

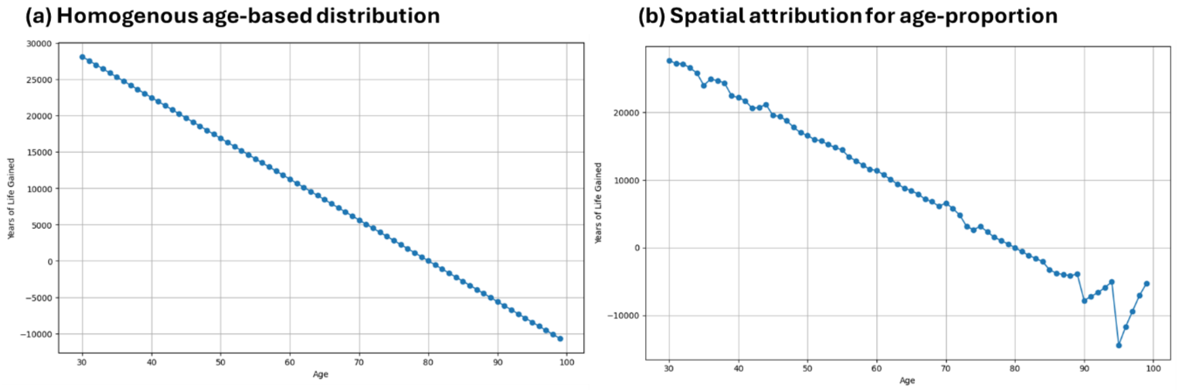


**Figure AIII.1: Sensitivity Analysis of Baseline Commune-Level Data for Years of Life Gained (YLG)**

While the differences in results between the two approaches is minor, the study prioritizes spatial attribution of age proportions for enhanced accuracy. Homogeneous age distribution assumptions simplify national-level and departmental-level computations but overlook essential geographic and demographic variability inherent to communes and IRIS units. In practice, communes exhibit significant variability in factors like population density, age structure, and exposure levels to pollutants, necessitating spatial granularity. Spatial attribution accurately represents areas with higher concentrations of vulnerable populations (e.g., elderly individuals), ensuring precision in health impact estimation. Relying solely on homogeneous assumptions could introduce biases in demographic diversity and projections, particularly over large or diverse regions. By integrating heterogeneous spatial attribution, the analysis ensures health estimates are robust, scientifically accurate, and grounded in localized population structures, while aggregated results at the commune level remain interpretable and policy-relevant.

#### Annexure IV - Methodology for assigning concentrations to IRIS and aggregation to Commune level from spatial grids

Pollutant concentrations were assigned to IRIS units and aggregated to the commune level using geospatial overlay and population-weighted aggregation of high-resolution concentration grids. The approach integrated modeled air pollution data with official administrative geometries to ensure spatially consistent exposure estimates. High-resolution gridded concentrations for $NO_2$ *and* $PM_{2.5}$ were obtained from the CHIMERE model and INERIS baseline simulations. Spatial boundaries for infra-municipal statistical units (IRIS) and communes were derived from IGN/INSEE datasets.

##### Step 1: Assignment of Gridded Concentrations to IRIS Units

Each grid cell was spatially overlaid with IRIS polygons. For every grid cell–IRIS intersection, the proportional area overlap was computed as:

$$Weights = \frac{\text{Area of (Grid Cell} \cap \text{IRIS)}}{\text{Area of IRIS}}$$

Gridded concentration values were then assigned to IRIS units using these overlap proportions as weights, ensuring that partial coverage of grid cells was appropriately accounted for in IRIS-level concentration estimates.

##### Step 2: Aggregation to Commune Level

IRIS-level concentrations were aggregated to the commune level using population-weighted averaging. This weighting scheme ensures that IRIS units with higher population contribute proportionally more to commune-level exposure estimates. For each commune, the average concentration was calculated as:

$$Concentration_{commune} = \frac{\sum(Conc_{IRIS} \times Population_{IRIS})}{\sum Population_{IRIS}}$$

##### Step 3: Calibration Using INERIS Baseline Data

To ensure consistency across modeled scenarios, concentration fields were calibrated using INERIS baseline simulations for 2019. For each grid cell, scenario-specific concentration differences were computed and these deltas were applied during IRIS-level assignment to adjust modeled concentrations relative to the baseline.

##### Step 4: Computation of Exposure Metrics

For each IRIS unit, exposure metrics including mean concentration and mean concentration change (delta) were calculated as weighted averages of intersecting grid cells, where weights correspond to normalized intersection proportions.

This methodology enables consistent and population-weighted downscaling of high-resolution gridded air pollution data to IRIS units and their aggregation to the commune level, supporting robust spatial exposure assessment for epidemiological and health impact analyses.

#### Annexure V – Application of Cessation Lag in Risk Assessment

Reductions in air pollution exposure (e.g. $PM_{2.5}$ and $NO_2$) are expected to lower population health risks. However, the health gains from improved air quality do not occur instantaneously, epidemiological evidence, expert panels, and clinical studies all indicate that risk lag-effect exist, i.e., a cessation lag between pollutant reduction and the full population-wide realization of health benefits. This annexure lays out the implementation, sensitivity analysis, and implications of applying cessation lag models to the health risk assessment framework.

**Cessation Lag Weighting and Cumulative Fractions**

To model the distribution of health benefits, two distinct cessation lag weighting schemes are utilized, COMEAP and a linear approach. The COMEAP weights are structured according to expert consensus with a lag of 20 years for mortality, where 30% of the benefits occur in the first year, 50% are distributed equally across years 2 through 5, and the remaining 20% are spread evenly from year 6 to a terminal year. Conversely, the linear lag approach assumes a uniform distribution where benefits accumulate equally across the specified lag period. In both instances, weights are normalized to ensure the total sum equals 1. The cumulative realization of these benefits is quantified through a cumulative effect fraction. For any evaluation year $y$ relative to a base year $t_0$, this fraction is calculated by summing the lag weights $w_i$ up to the elapsed time.

**Adjustment of Relative Risk (RR) and Exposure Interpolation**

The relative risk is adjusted to reflect the progressive realization of health benefits. The adjusted relative risk is determined by scaling the full-effect risk increment by the cumulative fraction:

$$RR_{adjusted} = RR_{full\ effect} \times effect\ fractions\ (f_t)$$

To calculate for each year, pollutant exposure levels are first determined through linear interpolation between defined anchor years (e.g., 2019, 2030, 2050). This ensures a continuous exposure profile across the study period. The full-effect RR for a specific endpoint is then derived from the exposure-response relationship, change in pollutant concentration and is the increment associated with the standard coefficient. The adjustment factor was identified for each year as the ratio of the cumulative excess risk under a lag scenario vs under no lag scenario. This gives us a scaling factor that tells what proportion of the immediate (no-lag) benefit is realised over the time horizon.

**Table AIV-2: Cessation lag considered for air pollutants in health-economic analysis**

| Health outcome | Pollutant | Phase-in-windows (Lags) |
|---|---|---|
| All-cause mortality (30+) | $PM_{2.5}$ / $NO_2$ | 0-20 years |
| Lung Cancer (35+) | $PM_{2.5}$ | 0-20 years |
| COPD (40+) | $PM_{2.5}$ / $NO_2$ | 0-10 years |
| Asthma (0–17) | $PM_{2.5}$ / $NO_2$ | 0 year |
| Asthma (18–39) | $PM_{2.5}$ | 0 year |
| ALRI (0–12) per episode | $NO_2$ | 0 year |
| Stroke (35+) | $PM_{2.5}$ | 0-5 years |
| Myocardial Infarction (30+) | $PM_{2.5}$ | 0-5 years |
| Hypertension (18+) | $PM_{2.5}$ | 0-5 years |
| Type 2 Diabetes (45+) | $PM_{2.5}$ | 0-10 years |

**Sensitivity tests:** To ensure the robustness of the primary findings, a sensitivity analysis is conducted by systematically varying the assumptions regarding exposure trajectories and the temporal distribution of health benefits.

**1. Sensitivity of the model to Lag Length:** To check the sensitivity of the models to different lag structures, we implemented different lag durations (5, 10, 20, 35 years) delay the accrual of health benefits when pollution drops after 2019. The ***figure AIV.1*** presents a side-by-side comparison of the time course of adjusted relative risk (RR) for $PM_{2.5}$ and $NO_2$, in response to a linear decline in exposure after a baseline year (~2019/2020), under various lag assumptions using the COMEAP weighting structure. The comparison showed all curves overlap initially, then diverge during the exposure reduction phase. Shortly after baseline,

RR begins to decrease as exposure declines, but the rate of decrease is modulated by the lag duration. Short lags (e.g., 5 yrs, blue) show earlier and more rapid RR improvement. Longer lags (20 or 35 yrs, green/red) cause a slower decline, with full benefit realized only after much more time has passed. The no lag line leads the others slightly at first (fastest RR drop), but is closely approached within 10–20 years, especially for shorter lags (5–10 yrs). Even 20-year lags only show a ~2-year "delay" in realizing the same relative risk reduction as the no-lag case. With a linear, gradual exposure trajectory, risk reduction is also gradual. Because a sizable portion of the health benefit accrues even in the early years for all but the longest lag models (e.g., COMEAP, with 30% first year, 50% in years 2–5), the curves don't dramatically diverge except for artificially long lags. The use of a linear, slow (monotonic) change means annual differences are small, so spreading the health response over more years doesn't create large gaps between lagged/no-lag lines. If exposure dropped *very suddenly* (step function), lags would produce a much larger divergence from the "no lag" benefit early on. Furthermore, for chronic policy analysis (30+ years), the adjustment is usually <15%, but in short-run economic appraisals or where policy aims for *rapid* health improvement, omission of lag can overestimate near-term benefits

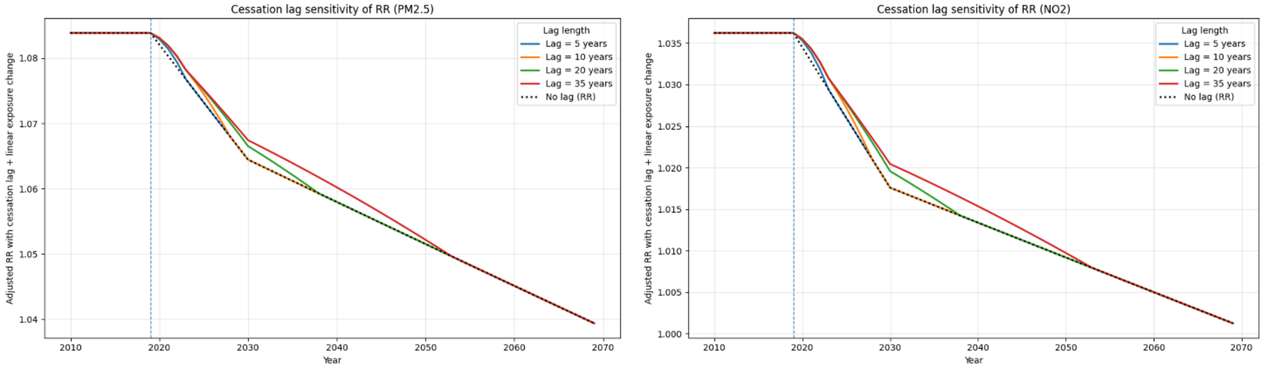


**Figure AIV.1 Sensitivity analysis of RR under multiple lag/trajectory assumptions**

**2. Sensitivity to linear exposure reduction & lag:** This figure AIV.2 presents a six-panel layout illustrating the application of cessation lag modeling to two pollutants $PM_{2.5}$ (left) and $NO_2$ (right), under the assumptions of a linear exposure reduction and a 20-year cessation lag using two lag models (COMEAP and linear), as well as a "no-lag" (immediate effect) comparator. For both $PM_{2.5}$ and $NO_2$, exposure drops steadily (~5–7 μg/m$^3$) between 2020 and 2070, with a monotonic decline. Linear interpolation between anchor years reflects an expectation of a gradual, well-managed rollout of policies. Linear exposure decline is a conservative policy assumption, it avoids overestimating early-year benefits that would result from a step-function (abrupt) exposure drop, which is rarely feasible in reality. It assumes a steady but achievable progression of air quality improvement, reflecting typical regulatory, technological, and behavioral implementation constraints. COMEAP is considered a biologically plausible and evidence-informed standard—it assumes some health benefits accrue quickly (early reversibility), but the majority are realized gradually, because underlying pathologies (e.g. chronic cardiovascular, respiratory) take years to resolve in the population. Linear lag offers an even more conservative approach, distributing benefits most slowly imaginable (uniform realization) and is likely to understate near-term gain. Using any lag at all is conservative compared to "no lag" (instantaneous), which is not epidemiologically credible for mortality or chronic disease endpoints.

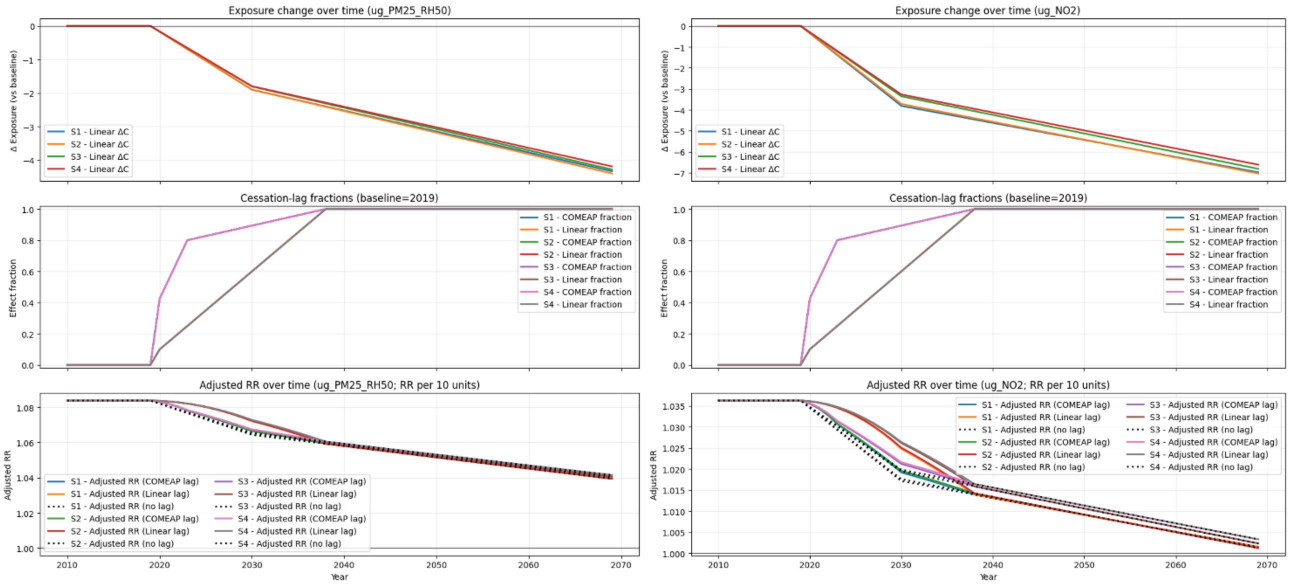


**Figure AIV.2: Comparison of linear exposure reductions, with COMEAP and linear lag fractions, and adjusted RR for $PM_{2.5}$ and $NO_2$ (20-year lag)**

**3. Sensitivity to non-linear exposure reduction & lag:** Both exposure and lag models are switched from linear to non-linear forms. Non-linear scenarios may mimic, for example, "slow start then rapid decrease" due to phased-in policies. The adjusted RR trajectory under non-linear exposure/lag models reveals that both the rate and timing of health gains can shift early benefits are smaller compared to linear but catch up closer to 2050. Blue dashed line shows "non-linear" (exponential) exposure trajectory, it stays flat before 2020 (deliberately kept constant), dips sharply around 2025–2035, and then flattens again. Orange dotted line depicts a "cosine" scenario, a wavy, smoother decline, representing a realistic situation like phased regulations or cyclical implementation. Both reduce total exposure by about 3.5 µg/m³, but the pace and timing of reductions differ. Non-linear/cosine models capture periods of slower and faster air quality improvement, which could reflect staged interventions or unexpected changes in policy/economy. However, it is important to note that the "no lag" versions for both non-linear and cosine trajectories almost perfectly overlap the lagged versions except for a slight delay in the inflection region. Therefore, the timing of health benefit realization (as seen in RR reduction) very closely follows the pattern of emission reduction. Lag effects are most pronounced where the exposure curve changes most rapidly. However, for both lag structures, there is only a slight temporal smoothing by 2050+, the RR reduction is nearly complete regardless of lag. Modeling both linear and non-linear pathways allows robust sensitivity analysis, helping policymakers understand when maximum health impacts can be expected and plan accordingly.

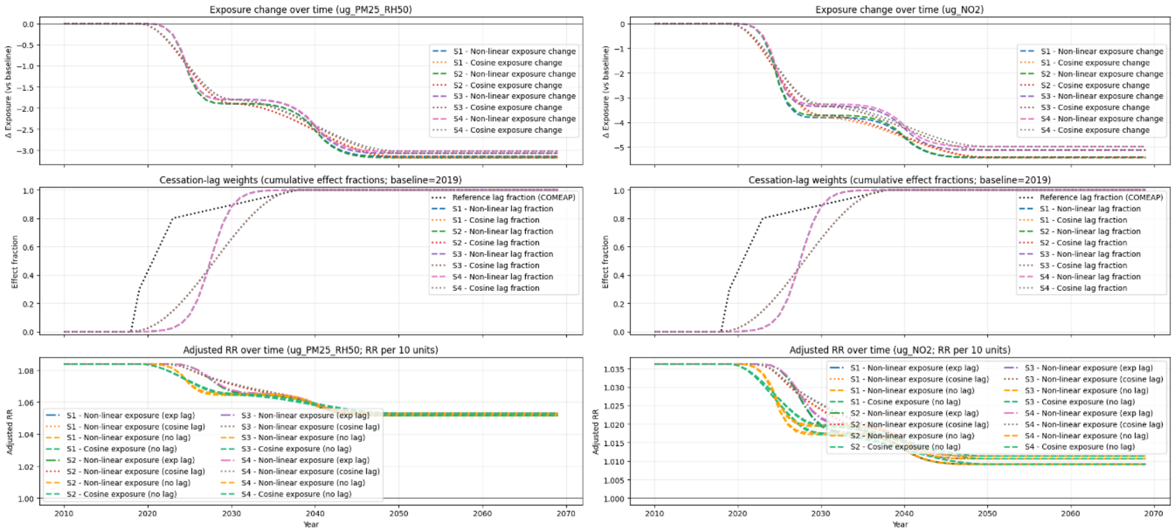


**Fig. AIV.3: Non-linear exposure trajectories and lag effects (dynamic policy scenario)**

#### Conclusion

In this analytic approach, the use of linear exposure interpolation and standardized, evidence-backed cessation lags (especially COMEAP), delivers a conservative and policy-relevant estimation of air quality health benefits over time. It ensures that predicted impacts are not overstated and remain robust under both clinical and implementation uncertainties. While pollutant concentrations in reality may fluctuate, adopting a linear approximation for exposure change between anchor years (e.g., 2019 to 2030) is a standard conservative baseline. In the absence of high-frequency policy data, assuming a steady linear decline avoids preempting exposure reductions, which could lead to an artificial inflation of health benefits in the early years of a projection. Furthermore, the use of the non-linear COMEAP framework is preferred over a simple linear lag for several reasons. A linear lag assumes that the 20$^{th}$ year after a policy change contributes exactly the same health benefit as the 1st year. This is rarely true in clinical practice. The COMEAP non-linear weights better reflect the stepped realization of health recovery. Both lag forms (COMEAP, linear) allowed us to explore the real-world impact of "*faster*" or "*slower*" benefit realization, informing scenario, uncertainty, and sensitivity analyses.
